# Linking GWAS to pharmacological treatments for psychiatric disorders

**DOI:** 10.1101/2022.05.11.22274981

**Authors:** Aurina Arnatkeviciute, Alex Fornito, Janette Tong, Ken Pang, Ben D. Fulcher, Mark A. Bellgrove

## Abstract

**Importance:** Large-scale genome-wide association studies (GWASs) are expected to inform the development of pharmacological treatments, however the mechanisms of correspondence between the genetic liability identified through GWASs and disease pathophysiology are not well understood.

**Objective:** To investigate whether functional information from a range of open bioinformatics datasets can elucidate the relationship between GWAS-identified genetic variation and the genes targeted by current treatments for psychiatric disorders.

**Design, Setting, Participants, and Exposures:** Relationships between GWAS-identified genetic variation and pharmacological treatment targets were assessed across four psychiatric disorders—ADHD, bipolar disorder, schizophrenia, and major depressive disorder. Using a candidate set of 2232 genes that are listed as targets for all approved treatments in the *DrugBank* database each gene was independently assigned two scores for each disorder – one based on its involvement as a treatment target, and the other based on the mapping between GWAS-implicated SNPs and genes according to one of four bioinformatic data modalities: SNP position, gene distance on the protein interaction network (PPI), brain eQTL, and gene expression patterns across the brain.

**Main Outcomes and Measures:** Gene scores for pharmacological treatments and GWAS-implicated genes were compared using a novel measure of weighted similarity applying a stringent null hypothesis-testing framework that quantified the specificity of the match by comparing identified associations for a particular disorder to a randomly selected set of treatments.

**Results:** Incorporating information derived from functional bioinformatics data in the form of PPI network revealed links for bipolar disorder (p^perm^ = 0.0001), however, the overall correspondence between treatment targets and GWAS-implicated genes in psychiatric disorders rarely exceeded null expectations. Exploratory analysis assessing the overlap between the GWAS-identified genetic architecture and treatment targets across disorders identified that most disorder pairs and mapping methods did not show a significant correspondence.

**Conclusions and Relevance:** The relatively low degree of correspondence across modalities suggests that the genetic architecture driving the risk for psychiatric disorders may be distinct from the pathophysiological mechanisms used for targeting symptom manifestations through pharmacological treatments and that novel approaches for understanding and treating psychiatric disorders may be required.

**Key Points:** *Question:* Do genes targeted by current treatments for psychiatric disorders match GWAS-identified genetic variation and what bioinformatic data modalities can inform these associations?

*Findings:* Information derived from functional bioinformatics data in the form of PPI network revealed links for bipolar disorder, however for most psychiatric disorders, the correspondence between GWAS-implicated genes and treatment targets did not exceed null expectations.

*Meaning:* GWAS-identified genetic variation driving the risk for psychiatric disorders may be distinct from the pathophysiological mechanisms influencing symptom onset and severity that are targeted by pharmacological treatments.

## Introduction

One of the core goals of genome-wide association studies (GWASs) is to inform the development of pharmacological treatments,^1^ under the assumption that if select genes are important in driving risk for a disorder, treatments that influence the function of these genes should mitigate risk and/or alleviate symptom expression. Current drugs for psychiatric disorders, however, were developed independently from these genomic findings, and it remains unclear whether the mechanisms of action for symptom reduction relate to GWAS identified genetic variation. In fact, whereas most pharmacological treatments in psychiatry work by modulating neurotransmission,^2–4^ psychiatric GWAS studies generally implicate genes involved in neurodevelopment, synaptic regulation, and plasticity.^5–7^ This apparent discrepancy may be a potential explanation for the relatively low efficacy of current psychiatric treatments,^4,8–12^ suggesting that genetic liability for the disorder might differ from the pathophysiological mechanisms that influence symptom onset and severity.

Converging evidence across complex disorders supports the overall relevance of genetic data in understanding and developing pharmacological treatments.^13–15^ For instance, drugs with genetic support move further along the development pipeline and are more likely to be clinically successful albeit the absolute size of the effect is relatively small.^16,17^ Moreover, genes that are less tolerant to mutations are more likely to be drug targets.^16^ GWAS-identified genes are also enriched in gene targets for drugs^18^ and demonstrate enrichment for relevant treatment-related gene categories such as antipsychotics relating to schizophrenia genes^19,20^ and antidepressants, anxiolytics, and antipsychotics linked to major depression genes.^21^ However, only a very small fraction of current drug targets for complex disorders can be identified through GWAS studies,^16,22–27^ indicating that the exact correspondence between GWAS findings and pharmacological treatments is limited.

Whereas the correspondence between GWAS findings and the respective treatment targets across a range of disorders is relatively low, incorporating functional information about gene action can provide a bridge between them. The main examples of integrating functional information to link pharmacological treatments and GWAS, however, come from studies of non-psychiatric disorders using connections in the protein interaction networks, Mendelian randomization approach, interactions through chromatin conformation, modulation of gene expression through eQTLs, or utilising the inverse relationship between the genetically-regulated and drug-induced expression signatures.^13–15,28–31^ Attempts to extend these functional genomics approaches to psychiatric disorders have been mainly limited to the analyses of transcriptomic data with the main focus on identifying potentially actionable treatment targets.^32–35^ Therefore, the utility of functional information derived from bioinformatic datasets to provide the link between the GWAS findings and corresponding treatments for psychiatric disorders remains unclear.

Here we introduce a novel method to evaluate the relationships between genes implicated in GWAS for psychiatric disorders and their pharmacological treatments according to several bioinformatics modalities using a stringent null hypothesis-testing framework. This allows us to determine the degree to which different sources of functional genomic information facilitate links between GWAS findings and drug targets; identify which functional data provide the most accurate correspondence; and assess whether treatments for phenotypically and genetically similar disorders show correspondence to their respective GWASs.

## Method

### Data availability statement

Data used in this manuscript are publicly available. Raw data files required for this project are hosted on a Figshare repository: https://doi.org/10.6084/m9.figshare.25356919. Full description of these datasets, code, and instructions for reproducibility are available at https://github.com/AurinaBMH/GWAS_drugs.

### Target selection

Main analyses focused on four psychiatric disorders: ADHD, schizophrenia, bipolar disorder, and major depressive disorder. We additionally used type 2 diabetes (T2D) as a non-psychiatric benchmark for comparison.^36^ To assess the correspondence between GWAS-implicated genes and drug targets, we curated a set of 2232 genes listed as targets for all approved treatments for all disorders in the *DrugBank* database^37^ [see details in eMethods 1 in Supplement 1]. For each disorder we selected a list of approved pharmacological treatments which included 14 drugs for ADHD, 29 for schizophrenia, 22 for bipolar disorder, 48 for major depressive disorder, and 45 for diabetes. Each treatment had a corresponding set of gene targets based *DrugBank* ^37^ (version 5.1.11), ranging from 63 gene targets (for ADHD) to 139 targets (for major depressive disorder) from the total set of 2232 [see details in eMethods 1 in Supplement 1]. We independently assigned each gene two scores for each disorder – one based on its involvement as a drug target for the disorder (as depicted in Fig. 1a), and the other based on the mapping between GWAS-implicated SNPs and genes according to one of four bioinformatic data modalities (as depicted in Fig.1 b-e).

**Figure 1.**
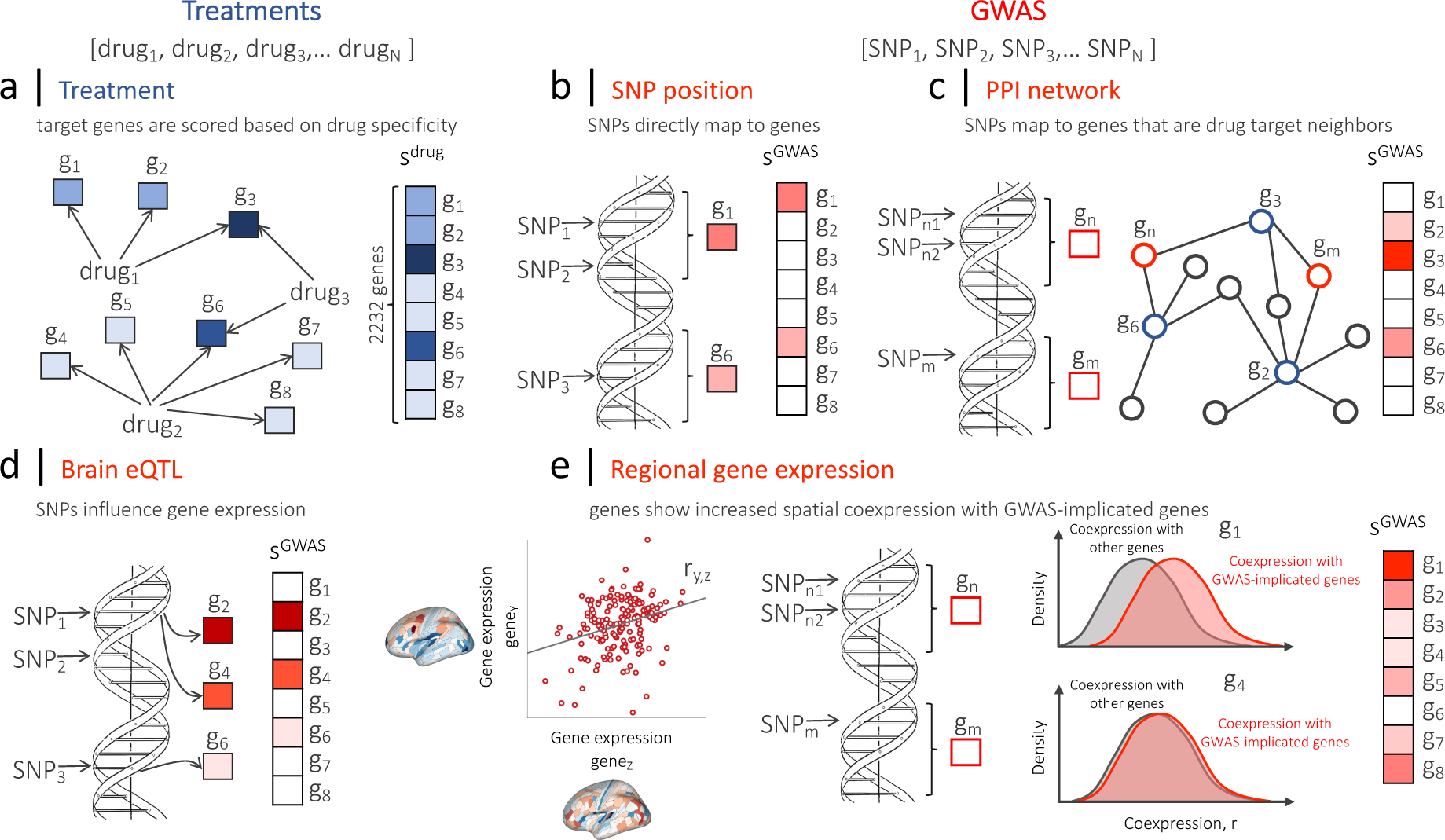
Evaluating the relationships between drug targets and GWAS by scoring genes based on: (i) their involvement in approved pharmaceutical treatments; and (ii) GWAS, for a given psychiatric disorder. (a) The analysis is based on a set of all clinically approved treatments. For each disorder, each of 2232 target genes derived from the DrugBank database^37^ are assigned scores corresponding to their role as a target across drugs approved for use in treating that disorder. A gene is scored based on its specificity for each drug by assigning a value of *w* = 1/L, where L is the total number of targets for that drug. The total score for a gene is quantified as a sum of scores across drugs. For each disorder, the same set of 2232 genes is also scored based on their similarity to variants implicated in corresponding GWASs using four main methods for mapping SNPs to genes: (b) SNP position, where the SNPs are mapped to genes based on their position on the DNA and the gene score consequently proportional to the cumulative effect size of all SNPs mapped to that gene; (c) PPI network, where genes are scored based on the proportion of their direct neighbors that are implicated in GWAS (as mapped based on SNP position); (d) brain eQTL, where genes are scored based on the cumulative impact of GWAS-identified SNPs on influencing their expression; (e) regional gene-expression similarity calculated from the Allen Human Brain Atlas (AHBA).^38^ First, for each pair of genes in the AHBA, the similarity of their spatial gene-expression profiles is quantified using a measure of gene-gene coexpression. Then, GWAS-implicated genes are identified based on SNP position and for each of 2232 target genes, coexpression value distributions are compared between GWAS-implicated and all other genes. As a result, a gene receives a high score if its coexpression with GWAS-implicated genes is, on average, higher than its coexpression with all other genes.

### Treatment-based scoring

Each gene was scored according to the specificity of each drug, where genes targeted by high-specificity drugs were assigned proportionally higher values [see eMethods 2 in Supplement 1]. Gene weights across all drugs for a disorder were then summed, producing a single treatment-based score vector, *s*^drug^, that independently measured the involvement of each gene as a treatment target for that disorder (Fig.1 a). As a result, genes that are targeted by more drugs with high specificity were assigned higher values compared to genes that are targets for fewer drugs or drugs with low specificity [see eMethods 2 in Supplement 1]. Genes that do not act as targets for that disorder were assigned a score of zero and consequently did not contribute to the similarity between GWAS-implicated genes and treatment targets.

### GWAS-based scoring

The GWAS-based score, denoted *s*^GWAS^, quantified a gene’s correspondence to GWAS identified variants, where similarity was defined with respect to one of four definitions (Fig. 1 b-e) that capture a distinct biological mechanism through which SNPs identified in a GWAS might influence genes: (i) the base-pair distance from the SNP on the DNA where genes with the highest number of significant SNPs get higher scores (Fig. 1 b); (ii) its topological proximity to the GWAS-implicated genes in the PPI network where we investigated the direct neighbours of those 2232 genes in the PPI network and calculated the proportion of these neighbours that were implicated in a GWAS (Fig. 1 c); (iii) its brain eQTLs quantifying the extent to which GWAS-identified SNPs act as eQTLs in brain tissue (Fig. 1 d); and (iv) its spatial pattern of gene expression across the brain^38^, where higher scores are assigned to genes whose expression patterns were more strongly correlated to genes identified through GWAS (Fig.1 e) [for all details see eMethods 3 in Supplement 1].

### Gene-score similarity

After scoring genes both according to their involvement in current disorder treatments (*s*^drug^) and to their implication by GWAS (*s*^GWAS^), we developed a method to assess the correspondence between the two sets of scores. We first normalized each of the score vectors across genes and calculated the inner product between these two unit-normalized vectors, resulting in a *weighted similarity score*, ρ [see eMethods 4 in Supplement 1]. As a result, only genes that were assigned non-zero scores for both *s*^drug^ and *s*^GWAS^ contributed to this similarity score.

### Evaluating significance based on random treatments

In contrast to previously used approaches that have applied relatively lenient significance testing (e.g. comparing a set of GWAS-implicated genes to all available genes),^16,22,23,39^ we evaluated the significance of a given weighted similarity score, ρ_emp_, using a permutation test comparing it to an ensemble of 5000 null values, ρ_rand_, generated by independently selecting a set of random drugs (while preserving the number of drugs associated with the disorder) [see eMethods 5 in Supplement 1]. As a result, a significant association indicates that GWAS-implicated genes match treatment targets for a selected disorder over and above what is expected by a random set of currently approved drugs as defined in the *DrugBank* database.

## Results

### Correspondence between GWAS-implicated genes and treatment targets

We first investigated whether current treatments for a given psychiatric disorder have gene targets that are more similar to their corresponding GWAS than a random set of drugs for each of the four mapping methods separately (Fig. 1 b-e). Mapping GWAS results to genes via the PPI network identified a significant correspondence for both diabetes (*p*^perm^ = 0.0001) and bipolar disorder (*p*^perm^ = 0.0001) (Fig. 2 b, permutation test against randomly selected drugs, statistically significant following Bonferroni correction for *n* = 5 tests, plotted as −log_10_(*p*) [see eMethods 5 in Supplement 1]), but no associations for schizophrenia, ADHD, or major depression.

**Figure 2.**
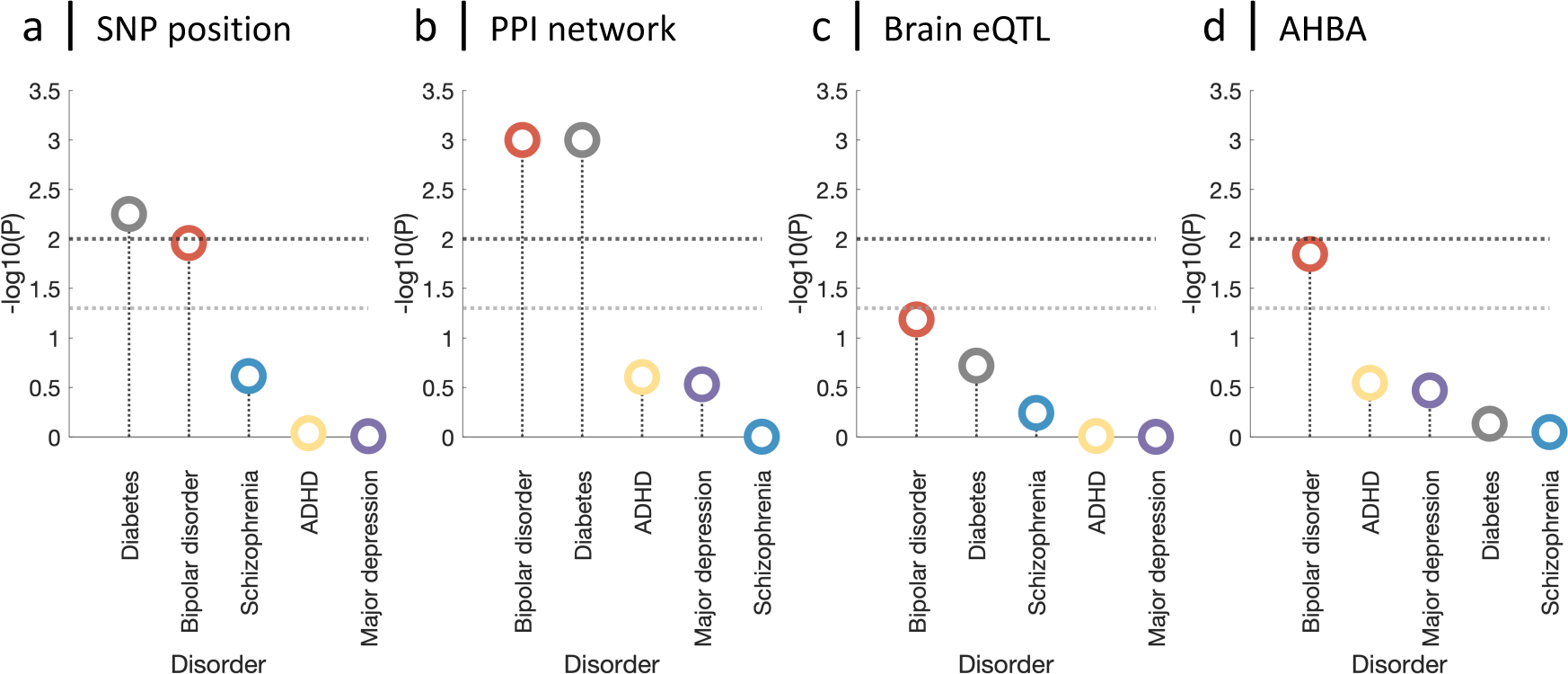
Bipolar disorder and diabetes demonstrate correspondence between treatment targets and genes implicated in GWAS data for each disorder. For a given disorder and mapping method, circles represent the significance of the association between the gene scores obtained from a set of drugs, s^drug^, and GWAS data, s^GWAS^, computed as a permutation test relative to a set of 5000 null scores generated from random drugs and plotted as - log_10_(p) [see details in eMethods 5 in Supplement 1]. Subplots correspond to the four GWAS mapping methods: (a) SNP position, (b) PPI network, (c) Brain eQTL, (d) Gene-expression similarity (cf. Fig. 1 b-e). Higher −log_10_(p) values indicate a stronger correspondence between GWAS and drug targets for a given disorder than expected from random treatments. Circles are colored by the disorder they represent (as labeled). Horizontal lines represent p-value thresholds: dark gray line – p = 0.01 (significant at α= 0.05 after Bonferroni correction for five disorders); light gray line – p = 0.05.

To shed light on the potential reasons why bipolar disorder was the only psychiatric disorder that displayed a GWAS-drug association, we performed a gene ontology (GO) enrichment analysis on both treatment and GWAS-based scores [see eMethods 6 in Supplement 1]. It indicated that treatment targets for all four psychiatric disorders are consistently enriched for GO categories related to synaptic signalling [see Supplement 2]. In contrast, the number of synaptic signalling-related categories for GWAS-implicated genes based on the PPI mapping was very low for all disorders except for bipolar disorder, where 9 of the top 20 categories involved transmembrane transport and other synaptic transmission and plasticity-related processes. These results suggest that the correspondence between treatment targets and GWAS-implicated genes for bipolar disorder is likely to occur due to the functional role of GWAS-implicated genes rather than the unique targets of bipolar disorder drugs.

For positional mapping the correspondence between drug targets and GWAS genes also significantly exceeded null expectation for diabetes (Fig. 2 a, *p*^perm^ = 0.0056). Whereas bipolar disorder demonstrated a relatively strong (*p*^perm^ < 0.05), but non-significant correspondence (*p*^perm^ > 0.01) using both positional and regional gene-expression-based mapping (Fig. 2 a,d). Associations for brain eQTL-based mapping did not exceed null expectation for any disorder (Fig. 2 c).

Our method also allows us to quantify the contribution of individual genes to the correspondence between GWAS and treatment-based scores, and thus delineate which genes drive the identified associations. The contribution of each gene to the similarity score, ρ, was quantified by comparing the inner product of 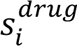 and 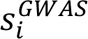 in real data to the distribution of inner product values derived from a null model [see eMethods 5 in Supplement 1]. To understand the functional roles of the most strongly implicated genes, we next investigated the associations identified for bipolar disorder and diabetes via PPI. We found that genes with the strongest involvement for bipolar disorder play roles in neurotransmission (*ADRA2C*, *GRIA3*, *GSK3B*, *HTR5A*, *HTR6*, *HTR7*, *p*^corr^ < 2.2 x 10^-5^, Bonferroni correction for *n* = 2232 tests), whereas genes implicated in diabetes were predominantly involved in insulin secretion and glucose metabolism (e.g., *ABCC8*, *DPP4*, *GLP1R*, *p*^corr^ < 2.2 x 10^-5^, Bonferroni correction for *n* = 2232 tests) [see eMethods 4 and eTable 1 in Supplement 1]. The fact that we identified genes with distinct and specific functional roles for both disorders was partially predetermined by their treatment targets, however, these genes contributed to the similarity between GWAS and treatment targets over and above the expectation based on the null model indicating their specific contribution.

We next performed exploratory analyses to assess whether treatments for some psychiatric disorders overlap with the GWAS genes of phenotypically and genetically similar disorders, expanding the comparison to all pairwise combinations of GWAS and treatment scores. Except for the previously identified associations for bipolar disorder and diabetes (eFigure 1 b (IV,V), d (IV)), most other disorder pairs and mapping methods did not show a significant correspondence after correcting for multiple comparisons. The strongest associations were identified for bipolar disorder, major depression, schizophrenia, and ADHD – treatment targets for bipolar disorder were linked to genes implicated in major depression (*p*^perm^ < 0.01), ADHD (*p*^perm^ < 0.01), and schizophrenia (0.01 < *p*^perm^ < 0.05) (eFigure 1 a (II, III), b (I, II, III), d (II, IV)), whereas treatment targets for schizophrenia were related to major depression and ADHD GWAS genes 0.01 < *p*^perm^ < 0.05) (eFigure 1 a (II), b (I)). Although not statistically significant, these cross-disorder associations are consistent with well-described phenotypic and genetic overlaps of schizophrenia, bipolar disorder, major depression, and ADHD.^40^

### The impact of data-processing choices

To identify whether different data-processing choices could improve the correspondence between GWAS and current treatment targets, we explored additional approaches for GWAS-to-drug target mapping such as representing the PPI network at varying densities, expanding eQTLs to include other tissues, as well as using data quantifying long-range chromatin interactions (Hi-c) resulting in a set of 27 measures [see eMethods 7 in Supplement 1]. As shown in eFigure 2, gene scores derived from different mapping methods demonstrated varying levels of similarity – PPI-based measures were generally more similar to each other than to Hi-c or eQTL-based scores, whereas regional gene expression was not similar to other measures.

The degree of correspondence between GWAS-implicated genes and treatment targets was markedly variable (Fig. 3): none of the individual mapping methods yielded significant matches for ADHD or schizophrenia (Fig. 3 a-c), whereas major depression, bipolar disorder, and diabetes exhibited significant correspondences for several PPI-based measures (Fig. 3 b,d,e). Consistent with the initial findings, gene scores assigned from the PPI network were generally the most informative for bipolar disorder, major depression, and diabetes, suggesting that indirect interactions through gene products might be involved in the mechanism of pharmacological treatments.

**Figure 3.**
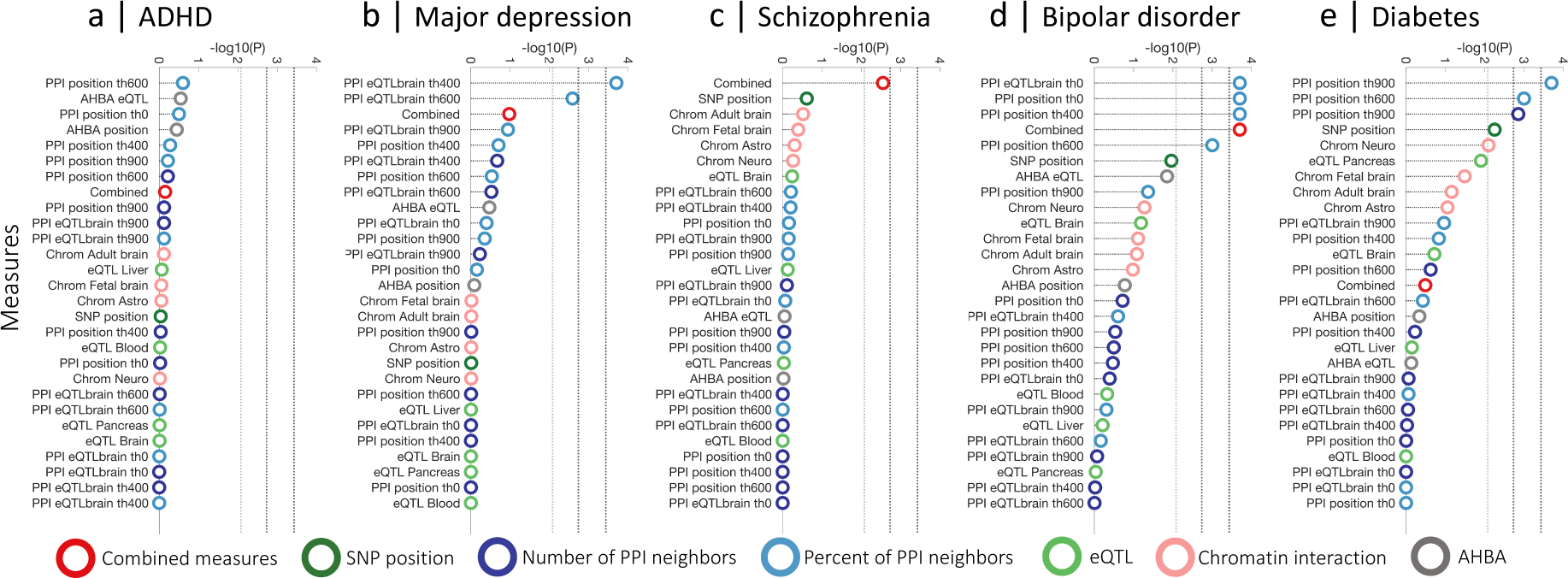
PPI-based mapping methods show the strongest correspondence between treatment targets and genes implicated in a GWAS for bipolar disorder and diabetes. Circles represent the significance of the association between the treatment-based score, s^drug^, and GWAS-based score, s^GWAS^, for a selected disorder. The significance is quantified using a permutation-based p-value comparing the empirical GWAS-treatment matching score ρ to a set of 5000 matching scores generated using a selection of random drugs [see details in eMethods 5 in Supplement 1]. The position of the circle corresponds to −log_10_(p) where higher values indicate a more significant association. Subplots correspond to different disorders: (a) ADHD, (b) major depression, (c) schizophrenia, (d) bipolar disorder, and (e) diabetes. Colors indicate different types of mapping methods: SNP position (dark green); spatial transcriptomic similarity (gray); chromatin interaction, Hi-c (pink); PPI network quantified as the proportion of neighbors (light blue); PPI network quantified as the total number of neighbors (dark blue); and eQTL (light green); linear combination of all measures (red; the null model was also adjusted to incorporate the linear regression across all measures in each iteration to account for this optimization step). Horizontal lines represent the degree of statistical significance: light gray line – p^perm^ = 8.3 × 10^-3^ (Bonferroni correction for six types of measures, this threshold provides guidance for the expected significance level considering that groups of measures derived from the same type of mapping method show a degree of similarity); dark gray line -- p^perm^ = 1.9 ×10^-3^ (Bonferroni correction for 27 measures); black line -- p^perm^ = 3.7 × 10^-4^ (Bonferroni correction for 27 measures and 5 disorders, this threshold provides guidance for a highly conservative correction assuming independence among all mapping methods and disorders). Permutation-based approach estimates a p-value with a minimum resolution of 2 × 10^-4^ (corresponding to 1/5000). If the estimated p-value is lower, we conservatively place it at 2 × 10^-4^.

We additionally evaluated whether complementary information derived from different mapping methods could increase correspondence to treatment targets. A linear combination of 27 available measures provided a statistically significant correspondence for bipolar disorder (*p*^perm^ = 2 × 10^-4^), and an improvement relative to all other measures for schizophrenia (*p*^perm^ = 0.003, slightly above the Bonferroni-corrected threshold for 27 measures, p = 0.002), but did not outperform individual measures for ADHD, major depressive disorder, and diabetes (Fig. 3). Whereas our results are mixed, the example of schizophrenia demonstrates that complementary information from multiple bioinformatic sources can sometimes be combined to substantially improve the correspondence between GWAS and treatment targets.

Overall similar results were obtained using different sets of GWAS summary statistics data for all psychiatric disorders^6,7,41,42^ as well as diabetes^43^ (eFigure 3-5) indicating that the results are not unduly influenced by the GWAS sample size. To assess if our findings were specific to psychiatric disorders, we also evaluated the correspondence between treatment targets and genes implicated by GWAS for several non-psychiatric diseases such as heart failure, rheumatoid arthritis, and inflammatory bowel disease. In most cases, individual mapping methods, as well as the combined score, did not provide significant correspondence between GWAS-implicated genes and treatment targets, suggesting poor mapping for these non-psychiatric disorders (eFigure 6).

## Discussion

In addition to identifying the genetic architecture of complex disorders, GWAS research aims to inform potential treatment targets that can be used in pharmacological interventions to alleviate the symptom expression. The possibility that risk genes for psychiatric disorders may act by creating vulnerability to illness during development through pathways that are distinct from the pathophysiological mechanisms influencing symptom onset and severity, however, is commonly overlooked. Here we aimed to investigate whether the mechanisms of action of pharmacological treatments specifically relate to the disorder-associated genetic variation identified through GWAS. To test this, we introduced a new method for assessing the correspondence between genetic variation implicated in GWAS for complex disorders and their pharmacological treatment targets using multiple bioinformatic modalities.

Whereas our method was able to identify associations for diabetes indicating its general utility for establishing meaningful links, in line with previous findings^16,22–27^ we found that the overall degree of correspondence between GWAS-implicated genes for psychiatric disorders and their respective treatment targets was low – only bipolar disorder exhibited significant associations, that were detectable when incorporating functional information derived from the PPI network. Importantly, these results were not strongly dependent on the size of the corresponding GWASs used in the analyses (eFigure 3-5), indicating that any lack of association does not simply arise from insufficient gene discovery power.

Our comprehensive analyses incorporating a range of bioinformatic modalities suggest that the genetic liability to psychiatric disorders might differ from the systems that are currently targeted by their pharmacological treatments. This implies that current treatments for these disorders are not designed to address the underlying causative events that lead to the development of the disorder. This notion is also supported by the fact that a large fraction of genes identified through GWASs relate to neurodevelopmental processes such as presynaptic and neuron differentiation, neuronal morphogenesis and projection,^5^ neuronal development and synapse formation,^6,44^ as well as synaptic plasticity,^7,41^ whereas targets for pharmacological treatments mostly involve genes for synaptic signalling indicating a likely disconnect between the genetic susceptibility and the mechanisms targeted by drugs.

Our results demonstrate that, among different data-processing options, PPI network-based mapping methods consistently yielded the strongest GWAS–drug associations (Fig. 3 b,d,e), supporting the idea that the concordance between GWAS and pharmacological treatments is likely to involve interactions between the corresponding gene products.^22,39,45^ These findings are also in line with the comprehensive evaluation of approaches for drug target identification demonstrating that incorporating diffusion through PPI network significantly increases drug target identification.^15^

Besides investigating the concordance between GWAS and treatment targets for individual disorders, our approach also allowed us to directly test cross-disorder correspondence – namely, whether treatment targets for one disorder match genes implicated in the GWAS of others. The suggestive associations identified between treatments for bipolar disorder and genes implicated in both major depression and schizophrenia are consistent with the general profile of bipolar disorder treatments as they target some of the overlapping symptoms between these disorders, such as depressive mood and psychotic episodes. Similarity in the genetic architecture between bipolar disorder, major depression, ADHD, and schizophrenia, quantified through the genetic correlation^40^ and common biological pathways,^46^ is in line with the observed coupling between GWAS and treatment targets. These findings accord with the results from a drug repurposing study indicating that repurposing candidates for bipolar disorder included a number of antipsychotics and antidepressants, whereas antipsychotics were commonly found among top hits for major depression.^32^

In contrast to previous studies which mainly evaluated significance with respect to gene set and pathway enrichment,^16,19–21,47^ or compared the identified correspondences to what could be expected using all available genes,^22,23,39^ our approach introduces a more stringent null model for evaluating associations based on other available treatments. In other words, instead of evaluating the correspondence compared to the rest of the genome, our approach addresses the specificity of those associations in the context of all available treatments by quantifying if the GWAS-implicated genes for a given disorder match its treatment targets to a higher extent than other drugs. Based on this null model, GWAS-implicated genes for a particular disorder are expected to demonstrate a specific coupling with corresponding treatments exceeding the associations with treatments for unrelated disorders. Our proposed method also offers the advantage of tailoring the null model by expanding or restricting the range of treatment categories used in generating the null distribution. For instance, when focusing on psychiatric disorders, we can restrict the null model to psychiatric treatments or adjust their categories based on other criteria (eFigure 7). The key advantage of such a modification is the ability to refine the testing based on the relevance of selected treatments.

## Conclusions

We observed a relatively low degree of correspondence between treatment targets and GWAS-implicated genes, suggesting that the genetic architecture of risk for psychiatric disorders may be distinct from the pathophysiological mechanisms targeted by current pharmacological treatments. This work encourages the further development of novel approaches for understanding and treating psychiatric disorders. With improvements in quantifying eQTLs, as well as the development of spatially comprehensive time-resolved and cell-specific gene-expression atlases similar methods can be easily used to investigate other complex disorders and examine potential cross-disorder associations.

## Data Availability

Code and links to all data are available online at: https://github.com/AurinaBMH/GWAS_drugs

## Acknowledgments

We are thankful to iPSYCH (Ditte Demontis and Anders Børglum) and deCODE (G. Bragi Walters, Hreinn Stefansson and Kari Stefansson) for giving us access to the recent GWAS meta-analysis summary statistics of ADHD.^44^ AA is a recipient of the Australian Research Council Discovery Early Career Researcher Award (project number DE230100498) funded by the Australian Government. MAB is supported by a National Health and Medical Research Council (NHMRC) of Australia Investigator Grant Level 2 (ID: 2025415). This work was supported by an ARC Linkage Project (LP160101592). AF was supported by the NHMRC (ID:1050504) and Sylvia and Charles Viertel Charitable Foundation. BDF was supported by National Health and Medical Research Council Grant 1089718. Authors have no competing interests.

