## Supplement 1 for "Linking GWAS to pharmacological treatments for psychiatric disorders"

**eMethods 1. Target selection**

The list of genes used in all analyses is based on the combined set of all approved treatment targets from the *DrugBank* database https://go.drugbank.com, version 5.1.11, downloaded on the 26th of February 2024. We selected all unique target genes annotated to the human species, resulting in a set, $G$, containing 2232 genes. Each gene in $G$ was then assigned a score, $g$, based on different independent criteria in relation to pharmacological treatments or GWASs. Scores assigned to all genes in $G$ can be represented as a score vector, $s_i$ ($i = 1, 2, ..., |G|$). Below we describe procedures for generating an $s_i$ according to treatment and GWAS-based scoring.

**eMethods 2. Treatment-based scoring**

Treatments for different conditions of interest were selected by searching www.drugbank.com (accessed September 3, 2020). Specifically, drugs for each indication were searched in the *DrugBank* database using the following search terms: 'attention deficit' (for ADHD); 'schizophrenia' (for schizophrenia); 'bipolar' (for bipolar disorder) excluding 'bipolar depression'; 'major depression' (for major depression); 'diabetes' (for type 2 diabetes) excluding 'type I diabetes' and 'diabetes insipidus'; 'heart failure' (for heart failure); 'Crohn's' and 'ulcerative colitis' (for inflammatory bowel disease); and 'rheumatoid arthritis' (for rheumatoid arthritis).

Gene targets for each drug were assigned based on the *DrugBank* database (version 5.1.11). The curated list of pharmacological treatments includes: ADHD (14 drugs with 63 unique gene targets); schizophrenia (29 drugs, 81 gene targets); major depression (48 drugs, 139 gene targets); bipolar disorder (22 drugs, 123 gene targets); diabetes (45 drugs, 106 gene targets). Later, we extend the analysis to other non-psychiatric conditions such as heart failure (46 drugs, 88 gene targets), rheumatoid arthritis (58 drugs, 115 gene targets), and inflammatory bowel disease (26 drugs, 41 gene targets).

Using this information, we constructed a score vector, $s^{\text{drug}}$, for each disorder across 2232 genes that quantifies the involvement of each gene as a treatment target for that disorder. To ensure that each drug was weighted equally, genes were scored with respect to the total number of targets for that drug: if a drug has $L$ target genes, then the score for each gene is incremented by an amount $1/L$. As a result, if a drug is very specific and has only a few targets, each of those target genes will be assigned a relatively high score compared to a less specific drug targeting multiple genes. The total treatment-based score, $s_i$, for gene $i$, is then computed as the sum of weights across all drugs. Genes with higher scores, $s_i$, for a given disorder, are those for which more drugs target them with high specificity; conversely, scores are minimal (0), for genes that are not targeted by any drugs used for treating a given disorder.

**eMethods 3. GWAS-based scoring**

For each disorder GWAS [ADHD,[1] schizophrenia,[2] major depression,[3] bipolar disorder,[4] diabetes,[5] heart failure,[6] rheumatoid arthritis,[7] and inflammatory bowel disease,[8] we constructed a score vector, $s_i^{GWAS}$, across 2232 genes that quantified the similarity of each gene to variants identified in each GWAS. The similarity was defined based on four main bioinformatic datasets including SNP position on the DNA, protein interaction network (PPI), brain eQTL, and regional gene expression across the brain as quantified using Allen Human Brain Atlas (AHBA).[9] Later, we expanded GWAS-based mapping methods to include non-brain eQTLs, brain chromatin interaction profiles (Hi-c) and explored the effects of different data-processing options.

***SNP position-based GWAS scoring***

This method relies on the assumption that SNPs located within protein-coding regions of a gene can potentially affect that gene by changing its functional products whereas SNPs in non-coding regions or in linkage disequilibrium (LD) with a particular gene can be involved in its regulatory mechanisms[10,11] (in some cases the LD relationships can extend between coding and non-coding regions linking these different types of SNPs).

For a given disorder's GWAS, each SNP was mapped to a corresponding gene, based on its position on the DNA using MAGMA software package https://ctg.cncr.nl/software/magma.[11] Corresponding annotation file `MAGMAdefault.genes.annot` and reference panel (1000 Genomes European ancestry) for linkage disequilibrium (LD) calculation were downloaded from https://github.com/thewonlab/H-MAGMA. Gene analyses in MAGMA are based on a multiple linear principal components regression model where the gene $p$-values are computed using an $F$-test. A score vector $s^{\text{GWAS-position}}$ was constructed using $-\log_{10}$ transformed gene $p$-values for each of 2232 genes $s_i^{GWAS-position} = -\log_{10}(P_i)$. Therefore, genes with a more significant involvement in a GWAS were assigned higher values. Genes that were absent from the MAGMA output were assigned a 0 score.

***PPI-based GWAS scoring***

Proteins encoded by specific genes exert their function through interactions with one another, therefore, indirect relationships between genes can be identified using functional information such as PPI networks.[12] We developed a gene-scoring procedure to incorporate information derived from the protein--protein interaction (PPI) network by investigating the direct neighbours of those 2232 genes in the PPI network and calculated the proportion of these neighbours that were implicated in a GWAS, As a result, we could quantify the degree to which a particular gene is associated with the GWAS-implicated genes through functional interactions.

PPI data containing the full network of scored links between proteins `9606.protein.links.v12.0.txt` were downloaded from the STRING database (version 12.0) on the 26th of February 2024 https://string-db.org/cgi/download.pl?sessionId=a1fHJhN5R9Md&species_text=Homo+sapiens. Protein interactions were quantified using confidence scores ranging from 150 to 999, with higher values assigned to interactions with stronger evidence. Applying different thresholds to those confidence scores allows rendering binary PPI networks where interactions exceeding a selected threshold are considered present while others are treated as absent. We selected a range of confidence score thresholds—0, 400, 600, and 900—yielding four binary PPI networks with varying densities. These networks were transformed into PPI distance matrices that capture the shortest path between all pairs of nodes using `distance_bin` function from the Brain Connectivity Toolbox.[13] We then matched proteins to their corresponding genes through `biomaRt` package in `Bioconductor`. The main analyses here are performed using PPI distance matrix generated from relatively high-confidence interactions (>600) providing a balance between sensitivity and specificity. However, note that we investigate the effect of this threshold in ***The impact of data-processing choices*** section.

The construction of PPI-based GWAS scores relied on the selection of genes implicated in each GWAS. To identify a list of genes for each GWAS, we controlled the family-wise error at 0.05 using Bonferroni correction (by setting a gene $p$-value threshold equal to 0.05 divided by the number of identified genes in MAGMA analysis). The number of identified genes for psychiatric disorder GWASs ranged from 95 for ADHD to 922 genes for schizophrenia. Our PPI-based scoring method assigned each of 2232 target genes ($g_i$) a score according to its vicinity to $G_{\text{GWAS}}$ genes on the PPI network along paths of length $k$. We started the analyses considering one-step paths, $k$=1 and for each of 2232 target genes identified all genes that directly interact with gene $g_i$ ($k$=1) resulting in a gene set, $G_{\text{int}}$. A subset of these genes were then labeled as being implicated in GWAS, $G_{\text{GWAS}} \in G_{\text{int}}$. The absolute number of overlapping genes for each target $g_i$ $G_{\text{GWAS}} \in G_{\text{int}}$) is proportional to the total number of its neighbors. Therefore, we scored each gene as the proportion of their interacting neighbors that are implicated in GWAS: $s_i^{GWAS-PPI} = | G_{\text{GWAS}} |/| G_{\text{int}} |$. Repeating the process across all 2232 genes we constructed the full score vector, $s^{\text{GWAS-PPI}}$. Following this general procedure, a score vector can be computed at any given path-length, $k$ (all genes within $k$ steps on the PPI network are included in $G_{\text{int}}$). Exploratory analyses indicated that increasing $k$ above 1 significantly reduces the specificity of the results, therefore all PPI-based analyses presented in this manuscript consider only 1-step neighbors ($k$=1).

##### *eQTL-based GWAS scoring*

SNPs that influence the expression of one or more genes are referred to as expression quantitative trait loci (eQTL) and can be located in close proximity or at a distance from that gene. Therefore, evaluating the extent to which GWAS-identified SNPs act as eQTLs in brain tissue allows quantification of the functional impact of these SNPs on the genes of interest. Using eMAGMA we constructed eQTL-based gene scores ($s^{\text{GWAS-eQTL}}$) quantifying each of 2232 genes based on their involvement in tissue-specific gene expression. eMAGMA is a validated pipeline based on the original MAGMA software that uses tissue-specific SNP-gene associations to assign SNPs to genes based on their association with gene expression. The SNP-gene associations are then aggregated in a gene-based test while adjusting for linkage disequilibrium and correlated gene expression.[14] Our initial analyses were based on brain eQTL information derived from the psychENCODE database[15] and then expanded to other tissues including liver, whole blood, and pancreas with corresponding annotation files derived from GTEx database[16] (version 8). Similarly to position-based gene scoring, a score vector $s^{\text{GWAS-eQTL}}$ was constructed using -$\log_{10}$ transformed gene $p$-values for each of 2232 genes $s_i^{GWAS-eQTL}$ = -$\log_{10}(P_i)$. Genes that were absent from the eMAGMA output were assigned a score of 0.

#### *Regional expression-based GWAS scoring*

Considering that psychiatric disorders are associated with altered gene expression in the brain,[17] and assuming that spatial gene-expression patterns are related to the mechanism of drug action,[18] we also incorporated spatial gene-expression data derived from the Allen Human Brain Atlas (AHBA).[9] Each gene was scored according to its spatial expression similarity to GWAS-implicated genes, giving higher scores to genes whose expression patterns were more strongly correlated to genes identified through GWAS. We developed a gene-scoring procedure based on high spatial resolution gene-expression data, that assigns a score for each gene based on its spatial gene co-expression patterns.

The AHBA[9] provides high-resolution whole-brain gene expression data derived from six post-mortem donor brains. First, genes implicated in GWAS were selected using the same procedure as described for PPI-based scoring. Here we included 15,744 genes that passed our quality-control criteria[19] across 180 regions of the left cortical hemisphere. First, we calculated a gene--gene coexpression matrix between each pair of 15,744 genes that captures correlations in regional expression profiles. High scores in the coexpression matrix indicated that a pair of genes had similar spatial expression patterns across the cortex. Gene scores, $s_i^{GWAS-AHBA}$, for each of 2232 genes, $g_i$, were then calculated by comparing each gene's coexpression with genes implicated in GWAS *vs* all other genes using *z*-scores from a Wilcoxon rank-sum test. Therefore, high scores indicate that a gene has stronger coexpression with GWAS-implicated genes than other genes.

#### *Hi-c-based GWAS scoring*

H-MAGMA assigns non-coding SNPs to their cognate genes based on long-range chromatin interactions in human brain tissue across two developmental epochs and two brain cell types measured by Hi-C.[20] This technique identifies developmentally specific and cell-specific neurobiologically relevant genes. Using H-MAGMA software[20] (https://github.com/thewonlab/H-MAGMA) we constructed four chromatin interaction gene scores $s^{\text{GWAS-Hi-C}}$ based on foetal brain, adult brain, neuronal, and astrocytic brain Hi-C. Similarly to eQTL and position-based gene scoring, a score vector $s^{\text{GWAS-Hi-C}}$ was constructed using $-\log_{10}$ transformed gene *p*-values for each of 2232 genes, as $s_i^{GWAS-Hi-C} = -\log_{10}(P_i)$.

#### eMethods 4. Gene-score similarity

We aimed to evaluate the extent to which genes involved in pharmacological treatments for psychiatric disorders match genes implicated in their corresponding GWAS. To achieve this, we independently constructed two sets of scores quantifying the involvement of each gene in:

I. pharmacological treatments $s_i^{drug}$; and
II. GWAS $s_i^{GWAS}$ based on different bioinformatic datasets (e.g. $s^{\text{GWAS-position}}$, $s^{\text{GWAS-PPI}}$, $s^{\text{GWAS-eQTL}}$, $s^{\text{GWAS-AHBA}}$).

The similarity between two different score vectors was defined using a similarity function $\rho(s^{\text{drug}}, s^{\text{GWAS}})$. First, each score vector was normalized as $\hat{s}_i^{(x)} = s_i^{(x)}/\sum_i s_i^{(x)}$, such that normalized score vectors define positive mass across genes that sums to unity; hence both drug and GWAS-based scores have an equal contribution. We then defined the weighted similarity score, ρ, as a simple inner product between two unit vectors $\rho = \sum_i \hat{s}_i^{drug} \hat{s}_i^{GWAS}$. Gene-score vectors in gene space that point in the same direction receive a maximal score, ρ= 1; those that are orthogonal receive a minimal score, ρ= 0. For example, if pharmacological treatments for a disorder target the same genes that are implicated in GWAS, then the resulting normalized score vectors, $\hat{s}^{drug}$ and $\hat{s}^{GWAS}$, will place similar weight on similar genes and thus obtain a high weighted similarity score, ρ.

The contribution of each gene to the similarity score ρ was quantified by comparing the inner product of $s_i^{\text{drug}}$ and $s_i^{GWAS}$ in real data to the distribution of inner product values derived using a null model [see eMethods 5. ***Evaluating significance based on random treatments***]. The gene ranking was based on the significance of *p*-values derived from such permutation testing.

#### eMethods 5. Evaluating significance based on random treatments

We evaluated the statistical significance of our results by estimating a *p*-value using a permutation test comparing empirically derived weighted similarity scores, $\rho_{\text{emp}}$, to an ensemble of 5000 null similarity scores generated by selecting a set of random drugs, $\rho_{\text{rand}}$. Specifically, for each disorder, we repeatedly selected N random drugs (where N is the number of drugs in the curated list of pharmacological treatments for that disorder) from the total of 2055 drugs in the *DrugBank* database and generated an ensemble of gene-score vectors, $\hat{s}^{drug}_{rand}$. Random gene-score vectors were then normalized and compared to the real $\hat{s}^{GWAS}$ resulting in a distribution of $\rho_{rand}$ values used to derive $p$-values.

Under the assumption that treatments for psychiatric disorders are likely to target different sets of genes compared to drugs for non-psychiatric conditions, we also tested the significance using a constrained set of treatments relevant only to psychiatric disorders. In this case, the random treatments were selected from a set of 82 drugs used to treat four psychiatric conditions investigated in our analysis, namely ADHD, schizophrenia, major depression, and bipolar disorder. Each disorder had a different number of available treatments, therefore, to avoid the over-sampling of drugs for disorders that have more treatments, we selected drugs from a probability distribution that gave an equal chance for all disorder drugs to be selected. Specifically, for each drug, the probability of being selected was inversely proportional to the total number of drugs for that disorder (1/N). If a drug is used to treat more than one disorder, these scores were summed. Similarly to the general case, we then generated an assembly of 5000 gene score vectors based on random drug selection for each disorder and used them to compute the distributions of $\rho_{randpsych}$ values to derive $p$-values.

**eMethods 6. Gene-set enrichment analysis using over-representation analysis**

Over-representation analysis (ORA) examines if there are gene sets [e.g., annotated using Gene Ontology (GO)] within a selected list of genes that are statistically over-represented in that list. Here we used ORA to investigate which GO categories are enriched in treatment-based scores ($s^{drug}$) and PPI-based GWAS scores ($s^{GWAS}$) for psychiatric disorders. Considering that only the minority of all 2232 genes were assigned non-zero scores in both $s^{drug}$ and $s^{GWAS}$, we applied a threshold to retain all genes with non-zero scores while using the remaining genes as a reference list. Functional gene group analyses were performed using version 3.2 of ErmineJ software.[21] Gene ontology annotations were obtained from GEMMA[22] https://gemma.msl.ubc.ca/arrays/showArrayDesign.html?id=735 as `Generic_human_ncbiIds_noParents.an.txt.gz` on March 4, 2024 (last updated 2024-03-01). Gene Ontology terms and definitions were automatically downloaded by ErmineJ on March 4, 2024 as `go.obo` (data version 2024-01-17) and can be downloaded from http://release.geneontology.org/2024-01-17/ontology/index.html. We performed ORA on the thresholded treatment-based scores ($s^{drug}$) and PPI-based GWAS scores ($s^{GWAS\text{-}PPI}$) for each psychiatric disorder testing the biological process GO categories with 5 to 100 genes available. The resulting *p*-values were corrected across 3609 and 3766 GO categories for $s^{\mathrm{drug}}$ and $s^{\mathrm{GWAS}}$ respectively, controlling the false discovery rate (FDR) at 0.05 using the method of Benjamini and Hochberg.[23]

**eMethods 7. Different data processing choices**

We investigated five types of changes:

1. First, we examined the effect of representing the PPI network at varying densities, by differently thresholding the pairwise confidence scores of protein—protein interactions: from 0 (the network includes all possible interactions), to 400 (where weakest interactions are not included), to 600 (used above), to 900 (only the interactions with the strongest evidence are kept) [see eMethods 3. ***PPI-based GWAS scoring***].
2. Second, we investigated the effect of scoring genes based on the total number of interacting neighbors that are implicated in the GWAS (rather than normalizing by the number of neighbors and computing a proportion of neighbors, as above) [see eMethods 3. ***PPI-based GWAS scoring***].
3. Third, in addition to selecting the initial set of genes for regional gene expression and PPI-based analyses using SNP position-based mapping ('PPI position', 'AHBA position'), we also performed these analyses using the initial set of genes based on brain eQTL mapping for these methods ('PPI eQTL brain', 'AHBA eQTL').
4. Fourth, we expanded brain-based eQTLs above to include other tissues such as blood, liver, and pancreas (hypothesizing that these might boost associations with T2D).
5. Fifth, we introduced a novel mapping method based on long-range chromatin interactions (Hi-c)[24] in human brain tissues across two developmental time points, allowing us to identify developmentally specific genes [see eMethods 3. ***Hi-c-based GWAS scoring***].

These modifications resulted in a total of 27 methods of mapping from GWAS SNPs to gene scores: via SNP position (1 measure), PPI network (16 measures), eQTL (4 measures), chromatin interactions (4 measures), and spatial transcriptomics (2 measures).

**eTable 1. Scoring individual genes based on their contribution to the similarity score,** ρ. The contribution of each gene to the similarity score, ρ, was quantified by comparing the inner product of $s_i^{drug}$ and $s_i^{GWAS}$ in real data to the distribution of inner product values derived using 5000 similarity values generated using a selection of random drugs. Genes with low *p*-values contribute to the similarity score ρ exceeding the null expectation. $p^{corr} < 2.2 \times 10^{-5}$ corresponds to Bonferroni correction for n = 2232 tests.

| **Bipolar disorder** | | **Diabetes** | |
|---|---|---|---|
| **Gene** | $p^{corr}$ | Gene | $p^{corr}$ |
| **ADRA2C** | 0 | ABCC8 | 0 |
| **GRIA3** | 0 | DPP4 | 0 |
| **GSK3B** | 0 | GLP1R | 0 |
| **HTR5A** | 0 | INSR | 0.0012 |
| **HTR6** | 0 | IGF1R | 0.0014 |
| **HTR7** | 0 | SLC5A2 | 0.0024 |
| **DRD3** | 0.0002 | PPARG | 0.0044 |
| **DRD4** | 0.0002 | CPE | 0.0204 |
| **HTR2A** | 0.0004 | IGFBP7 | 0.0204 |
| **HTR1E** | 0.0018 | NPY | 0.0204 |
| **SCN11A** | 0.0028 | AMY2A | 0.0230 |
| **SCN8A** | 0.0028 | ETFDH | 0.0232 |
| **DRD2** | 0.0044 | GPD1 | 0.0232 |
| **HTR1A** | 0.0058 | PRKAB1 | 0.0234 |
| **CDKN1A** | 0.0090 | ABCB11 | 0.0236 |
| **HTR2C** | 0.0092 | TRPM4 | 0.0236 |
| **OGDH** | 0.0102 | GANAB | 0.0240 |
| **HGF** | 0.0108 | RAMP1 | 0.0240 |
| **HTR2B** | 0.0112 | RAMP2 | 0.0240 |
| **SCN3A** | 0.0142 | RAMP3 | 0.0240 |

**eFigure 1. Cross-disorder comparison.**

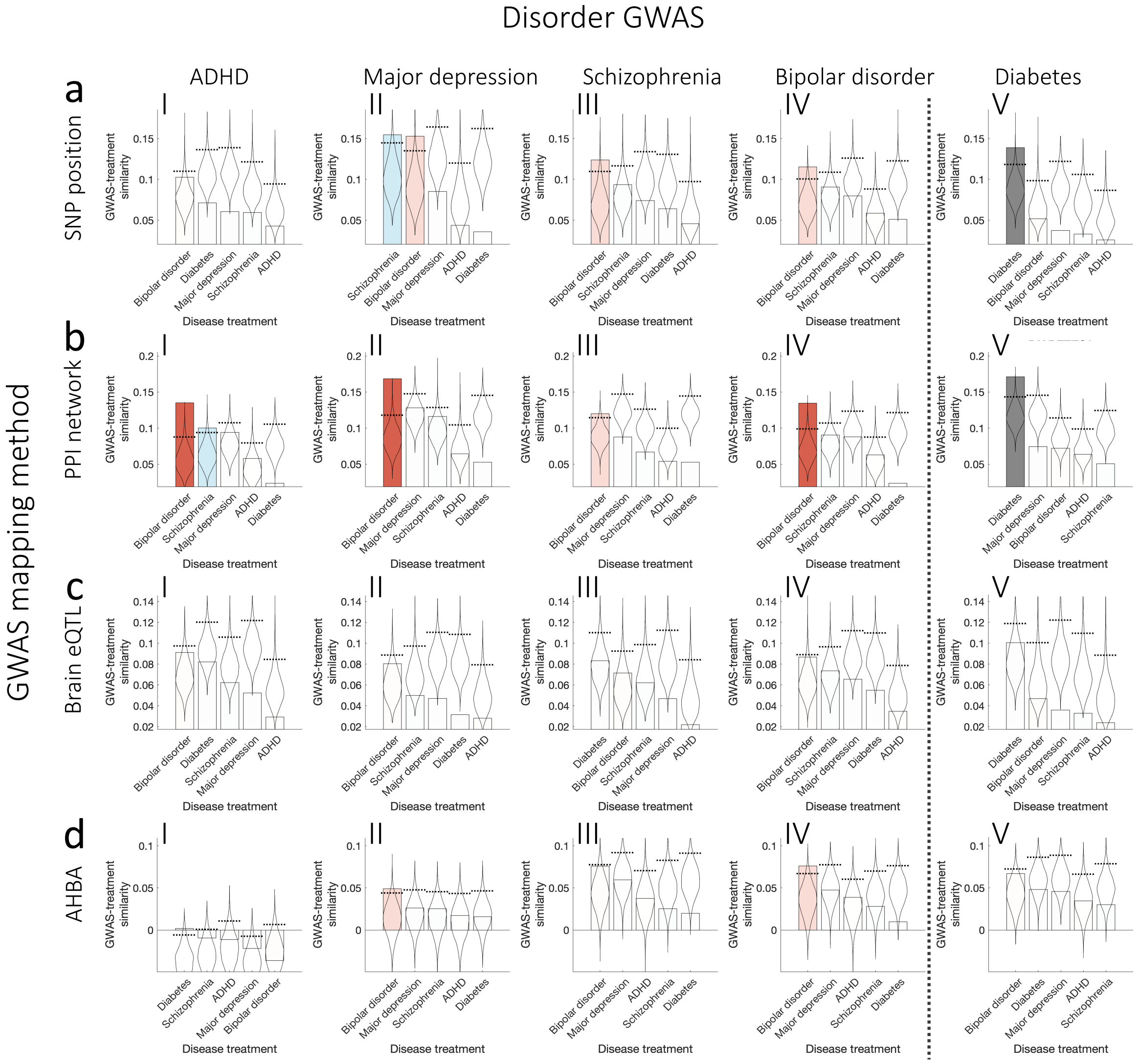


Note: **The pairwise correspondence between treatment targets and genes implicated in GWAS for each disorder.** Each column corresponds to a different disorder GWAS: ADHD, major depression, schizophrenia, bipolar disorder, and diabetes. Each row corresponds to a different GWAS mapping method: (a) SNP position, (b) PPI network, (c) Brain eQTL, (d) regional gene expression as quantified using AHBA. Subplots represent GWAS treatment similarity scores (ρ) for a selected pair of the mapping method and disorder GWAS for each disorder treatment list. Bars correspond to GWAS-treatment similarity values (ρ). Each bar has a corresponding null distribution presented in a form of a violin plot of 5000 similarity values generated using a selection of random drugs. The color of each bar corresponds to each treatment list: bipolar disorder (red), diabetes (gray), schizophrenia (blue), and ADHD (yellow). Color intensity indicates the statistical significance of the similarity score compared to the null distribution: dark represents p-values exceeding permutation testing $p^{perm} < 0.01$ (Bonferroni correction for five treatment lists); light represents $0.01 < p^{perm} < 0.05$; white represents $p^{perm} > 0.05$.

#### eFigure 2. Correlations between mapping methods.

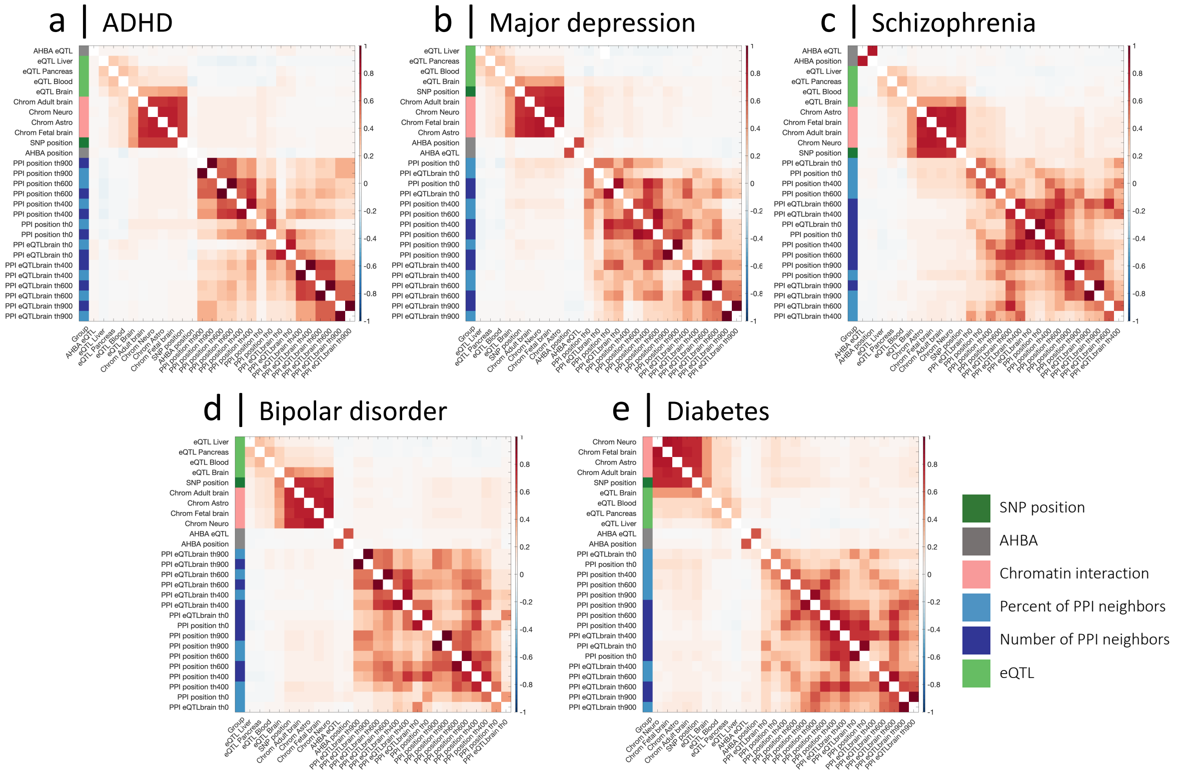


Note: **Correlation between mapping methods**. Each matrix represents the correlation between GWAS-based scores ($s^{GWAS}$) defined based on different mapping methods for each disorder: (a) ADHD, (b) major depression, (c) schizophrenia, (d) bipolar disorder, (e) diabetes. Similarity between each pair of $s^{GWAS}$ is quantified as a Spearman correlation. Colors on the left indicate different types of mapping methods: SNP position (dark green); spatial transcriptomic similarity (gray); chromatin interaction, Hi-c (pink); PPI network quantified as the proportion of neighbors (light blue); PPI network quantified as the total number of neighbors (dark blue); and eQTL (light green). PPI and chromatin interaction-based mapping methods are more similar to each other compared to other measures.

#### eFigure 3. Exploring different mapping methods for non-psychiatric disorders.

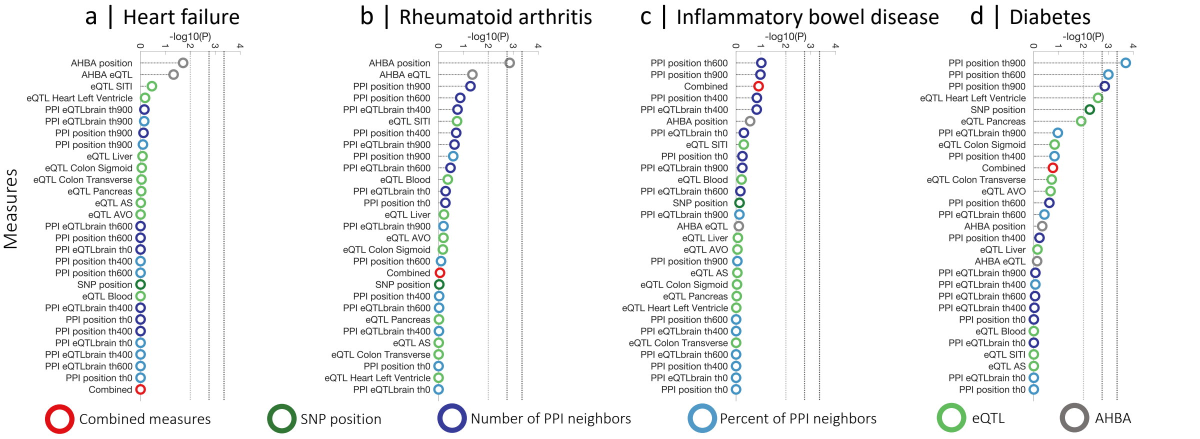


Note: **The correspondence between treatment targets and genes implicated in a GWAS for non-psychiatric disorders across different mapping methods.** Circles represent the significance of the association between the treatment-based score, $s^{drug}$, and GWAS-based score, $s^{GWAS}$, for a selected disorder. The significance is quantified using a permutation-based *p*-value comparing the empirical GWAS-treatment matching score ρ to a set of 5000 matching scores generated using a selection of random drugs. The position of the circle corresponds to −log10(*p*) where higher values indicate more significant association. Subplots correspond to different disorders: (a) heart failure, (b) rheumatoid arthritis, (c) inflammatory bowel disease, and (d) diabetes. Colors indicate different types of mapping methods: SNP position (dark green); spatial transcriptomic similarity (gray); PPI network quantified as the proportion of neighbors (light blue); PPI network quantified as the total number of neighbors (dark blue); and eQTL (light green); linear combination of all measures (red). Horizontal lines represent the degree of statistical significance: light gray line – $p^{perm} = 0.01$ (Bonferroni correction for five types of measures, this threshold provides guidance for the expected significance level considering that groups of measures derived from the same type of mapping method show a degree of similarity); dark gray line – $p^{perm} = 1.8 \times 10^{-3}$ (Bonferroni correction for 28 measures); black line – $p^{perm} = 4.4 \times 10^{-4}$ (Bonferroni correction for 28 measures and 4 disorders, this threshold provides guidance for a highly conservative correction assuming independence among all mapping methods and disorders). Permutation-based approach estimates a p-value with a minimum resolution of $2 \times 10^{-4}$ (corresponding to 1/5000). If the estimated *p*-value is lower, we conservatively place it at $2 \times 10^{-4}$.

### Replicating results using an alternative set of GWAS summary statistics.

For these analyses the following GWAS summary statistics were used: ADHD – Demontis et al.,[25] schizophrenia – Ripke et al.,[26] major depression – Howard et al.,[27] bipolar disorder – Ruderfer et al.,[28] diabetes – Xue et al.[29]

#### eFigure 4. The correspondence between treatment targets and genes implicated in GWAS data for each disorder.

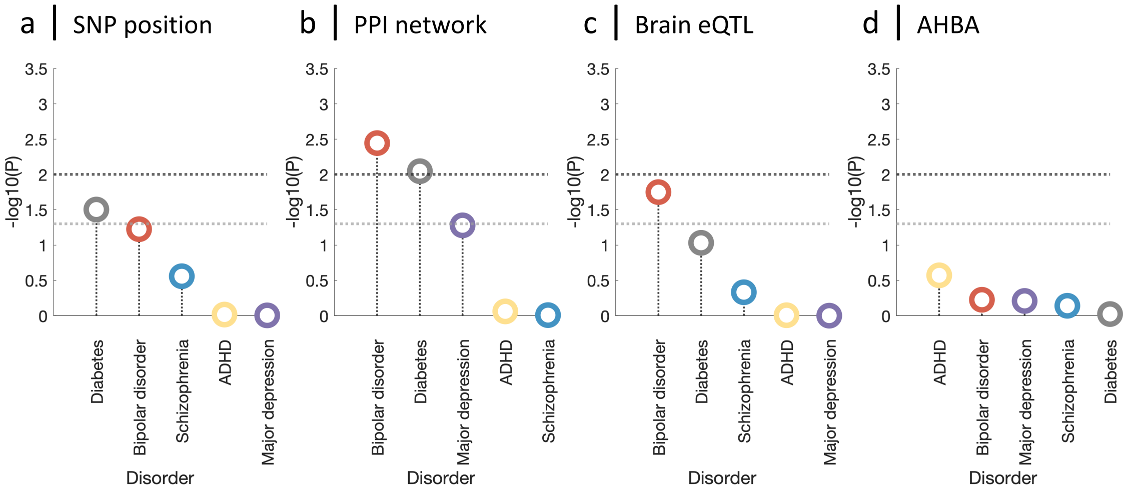


Note: **The correspondence between treatment targets and genes implicated in GWAS data for each disorder.** For a given disorder and mapping method, circles represent the significance of the association between the gene scores obtained from a set of drugs, $s^{drug}$, and GWAS data, $s^{GWAS}$, as $-\log 10(p)$. Subplots correspond to four GWAS mapping methods: (a) SNP position, (b) PPI network, (c) Brain eQTL, (d) Cortical gene-expression similarity. Statistical significance is quantified using a permutation-based approach in which the empirical GWAS-treatment matching score, ρ, is compared to a set of 5000 null scores generated from sets of random drugs. The position of each circle corresponds to $-\log 10(p)$; higher values indicate a more significant association between GWAS-based gene scores and the gene targets of treatments (relative to random treatments). The color of each circle corresponds to each disorder. Horizontal lines represent the degree of statistical significance: dark gray line – $p^{perm} = 0.01$ (Bonferroni correction for 5 disorders); light gray line – $p^{perm} = 0.05$.

**eFigure 5. The pairwise correspondence between treatment targets and genes implicated in GWAS for each disorder**.

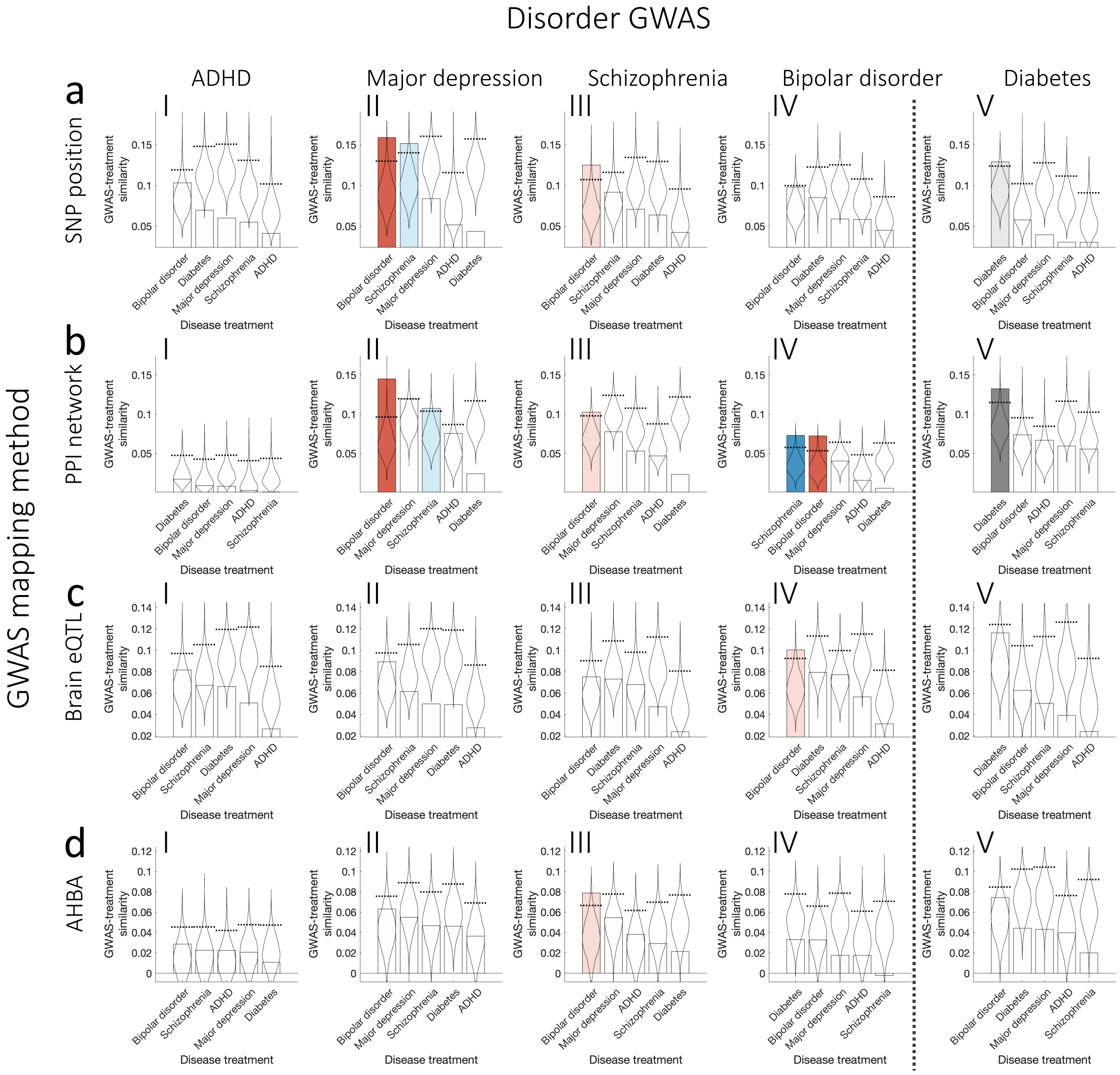


Note: **The pairwise correspondence between treatment targets and genes implicated in GWAS for each disorder.** Each column corresponds to a different disorder GWAS: ADHD, major depression, schizophrenia, bipolar disorder, and diabetes. Each row corresponds to a different GWAS mapping method: (a) SNP position, (b) PPI network, (c) Brain eQTL, (d) regional gene expression as quantified using AHBA. Subplots represent GWAS-treatment similarity scores (ρ) for a selected pair of the mapping method and disorder GWAS for each disorder treatment list. Bars correspond to GWAS-treatment similarity values (ρ). Each bar has a corresponding null distribution presented in a form of a violin plot of 5000 similarity values generated using a selection of random drugs. The color of each bar corresponds to each treatment list: bipolar disorder (red), diabetes (gray), and schizophrenia (blue). Color intensity indicates the statistical significance of the similarity score compared to the null distribution: dark represents *p*-values exceeding permutation testing $p^{perm} < 0.01$ (Bonferroni correction for five treatment lists); light represents $0.01 < p^{perm} < 0.05$; white represents $p^{perm} > 0.05$.

#### eFigure 6. The correspondence between treatment targets and genes implicated in a GWAS for each disorder across different mapping methods.

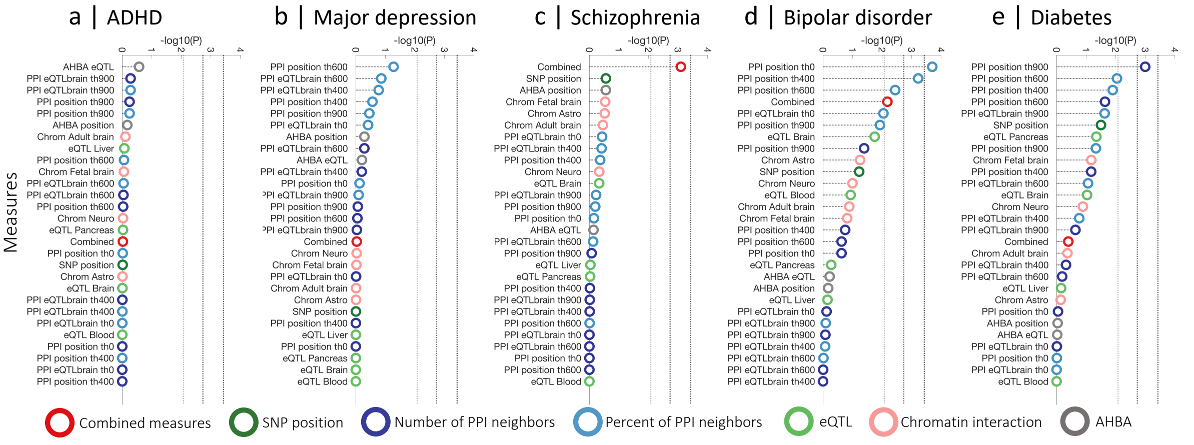


Note: **The correspondence between treatment targets and genes implicated in a GWAS for each disorder across different mapping methods.** Circles represent the significance of the association between the treatment-based score, $s^{drug}$, and GWAS-based score, $s^{GWAS}$, for a selected disorder. The significance is quantified using a permutation-based p-value comparing the empirical GWAS-treatment matching score ρ to a set of 5000 matching scores generated using a selection of random drugs. The position of the circle corresponds to $-\log10(p)$ where higher values indicate more significant association. Subplots correspond to different disorders: (a) ADHD, (b) major depression, (c) schizophrenia, (d) bipolar disorder and (e) diabetes. Colors indicate different types of mapping methods: SNP position (dark green); spatial transcriptomic similarity (gray); chromatin interaction, Hi-c (pink); PPI network quantified as the proportion of neighbors (light blue); PPI network quantified as the total number of neighbors (dark blue); and eQTL (light green); linear combination of all measures (red). Horizontal lines represent the degree of statistical significance: light gray line – $p^{perm} = 8.3 \times 10^{-3}$ (Bonferroni correction for six types of measures, this threshold provides guidance for the expected significance level considering that groups of measures derived from the same type of mapping method show a degree of similarity); dark gray line – $p^{perm} = 1.9 \times 10^{-3}$ (Bonferroni correction for 27 measures); black line – $p^{perm} = 3.7 \times 10^{-4}$ (Bonferroni correction for 27 measures and 5 disorders, this threshold provides guidance for a highly conservative correction assuming independence among all mapping methods and disorders). Permutation-based approach estimates a *p*-value with a minimum resolution of $2 \times 10^{-4}$ (corresponding to 1/5000). If the estimated *p*-value is lower, we conservatively place it at $2 \times 10^{-4}$.

**eFigure 7. Null models for significance testing**

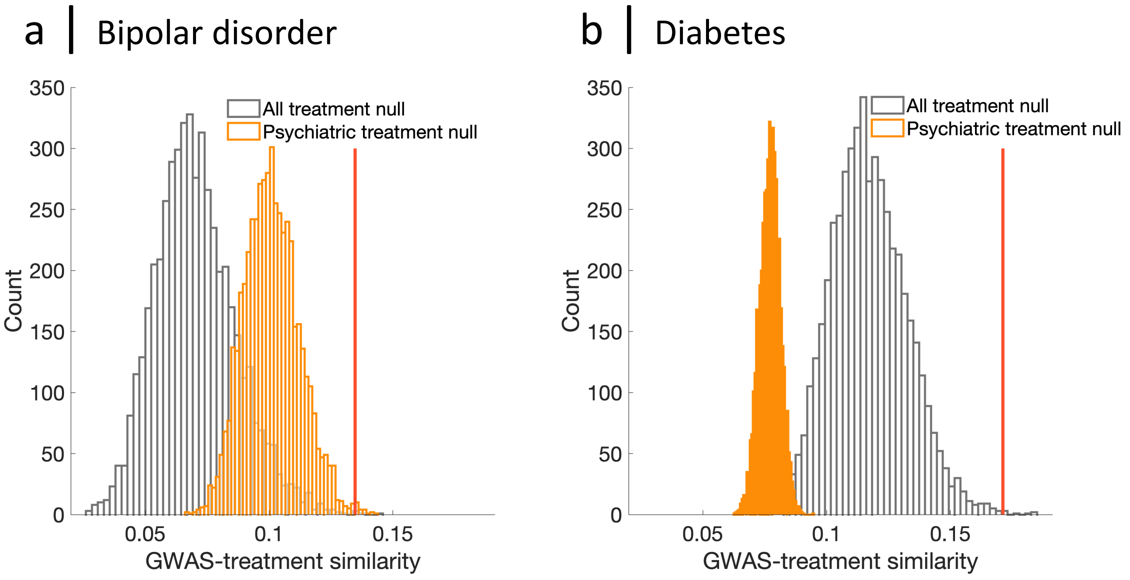


Note: **The comparison of different null models for significance testing.** Null distributions of the similarity scores used for evaluating the statistical significance of the correspondence between genes implicated by disorders GWAS and own treatment targets for (a) bipolar disorder and (b) diabetes. Distributions presented in gray are generated using sets of any random drugs ($\rho_{rand}$). Distributions presented in orange are generated using sets of random drugs for psychiatric disorders ($\rho_{randpsych}$). In each case, the vertical line indicates the empirical similarity value ($\rho$). The extent to which the significance of the association depends on the null model is related to the type of the disorder: bipolar disorder shows only small differences ($p_{rand} = 0.008$, vs $p_{randpsych} = 0.001$), whereas more considerable divergence is observed for diabetes ($p_{rand} = 0.003$, vs $p_{randpsych} = 0$). The null model based only on psychiatric drugs results in a much narrower null distribution modifying the significance of the association for both disorders. It is likely that genes driving the correspondence for bipolar disorder are generally more targeted by other psychiatric drugs and less similar to gene targets for other psychiatry unrelated treatments resulting in a right-shift of the distribution compared to the null based on all available drugs. On the contrary, drug targets for diabetes are generally less similar to genes implicated in psychiatric treatments, therefore, the null distribution in this case shows a left-shift increasing the significance of identified association. These differences highlight that the choice of a null model can modify the question addressed within the analysis.
