## Supplement 2 for "Linking GWAS to pharmacological treatments for psychiatric disorders"

**eTable 2. Gene enrichment results. Gene ontology biological processes categories implicated in ADHD treatment-based scores using over-representation analysis (false discovery rate (FDR) corrected p < 0.05).**

| | GOcategory | Description | NumGenes | Pval | Pval_corr |
|---|---|---|---|---|---|
| 1 | GO:0098664 | G protein-coupled serotonin receptor signaling pathway | 21 | 0 | 1.20E-14 |
| 2 | GO:0007187 | G protein-coupled receptor signaling pathway, coupled to cyclic nucleotide second messenger | 45 | 0 | 1.20E-14 |
| 3 | GO:0007188 | adenylate cyclase-modulating G protein-coupled receptor signaling pathway | 92 | 0 | 1.60E-14 |
| 4 | GO:0071880 | adenylate cyclase-activating adrenergic receptor signaling pathway | 12 | 0 | 3.20E-14 |
| 5 | GO:0071875 | adrenergic receptor signaling pathway | 13 | 0 | 1.65E-13 |
| 6 | GO:0035150 | regulation of tube size | 65 | 2.38E-13 | 1.07E-10 |
| 7 | GO:0035296 | regulation of tube diameter | 65 | 2.38E-13 | 1.07E-10 |
| 8 | GO:0097746 | blood vessel diameter maintenance | 65 | 2.38E-13 | 1.07E-10 |
| 9 | GO:0007193 | adenylate cyclase-inhibiting G protein-coupled receptor signaling pathway | 43 | 1.50E-12 | 6.01E-10 |
| 10 | GO:0007200 | phospholipase C-activating G protein-coupled receptor signaling pathway | 36 | 2.26E-12 | 8.14E-10 |
| 11 | GO:0003018 | vascular process in circulatory system | 82 | 1.48E-11 | 4.84E-09 |
| 12 | GO:0007197 | adenylate cyclase-inhibiting G protein-coupled acetylcholine receptor signaling pathway | 8 | 7.20E-11 | 2.00E-08 |
| 13 | GO:0007213 | G protein-coupled acetylcholine receptor signaling pathway | 8 | 7.20E-11 | 2.00E-08 |
| 14 | GO:0095500 | acetylcholine receptor signaling pathway | 12 | 1.03E-10 | 2.66E-08 |
| 15 | GO:0003013 | circulatory system process | 94 | 1.51E-10 | 3.64E-08 |
| 16 | GO:0090066 | regulation of anatomical structure size | 96 | 2.15E-10 | 4.86E-08 |
| 17 | GO:0098926 | postsynaptic signal transduction | 14 | 5.99E-10 | 1.27E-07 |
| 18 | GO:0042310 | vasoconstriction | 16 | 2.46E-09 | 4.92E-07 |
| 19 | GO:0007189 | adenylate cyclase-activating G protein-coupled receptor signaling pathway | 51 | 4.90E-09 | 9.30E-07 |
| 20 | GO:0060078 | regulation of postsynaptic membrane potential | 72 | 3.26E-08 | 5.89E-06 |
| 21 | GO:0006940 | regulation of smooth muscle contraction | 34 | 1.68E-07 | 2.89E-05 |
| 22 | GO:0007631 | feeding behavior | 27 | 3.33E-07 | 5.46E-05 |
| 23 | GO:0007626 | locomotory behavior | 61 | 4.40E-07 | 6.91E-05 |
| 24 | GO:0015872 | dopamine transport | 8 | 7.49E-07 | 1.13E-04 |
| 25 | GO:0015844 | monoamine transport | 14 | 9.22E-07 | 1.33E-04 |

**eTable 3. Gene enrichment results. Gene ontology biological processes categories implicated in ADHD GWAS-based scores using over-representation analysis (false discovery rate (FDR) corrected p < 0.05).**

| | GOcategory | Description | NumGenes | Pval | Pval_corr |
|---|---|---|---|---|---|
| 1 | GO:0016241 | regulation of macroautophagy | 50 | 1.60E-14 | 6.16E-11 |
| 2 | GO:0051896 | regulation of phosphatidylinositol 3-kinase/protein kinase B signal transduction | 73 | 2.27E-11 | 3.40E-08 |
| 3 | GO:0007035 | vacuolar acidification | 18 | 3.06E-11 | 3.40E-08 |
| 4 | GO:0010506 | regulation of autophagy | 88 | 3.62E-11 | 3.40E-08 |
| 5 | GO:0010564 | regulation of cell cycle process | 97 | 9.04E-11 | 5.52E-08 |
| 6 | GO:0097553 | calcium ion transmembrane import into cytosol | 48 | 9.05E-11 | 5.52E-08 |
| 7 | GO:0050731 | positive regulation of peptidyl-tyrosine phosphorylation | 57 | 1.18E-10 | 5.52E-08 |
| 8 | GO:0002181 | cytoplasmic translation | 20 | 1.19E-10 | 5.52E-08 |
| 9 | GO:0071900 | regulation of protein serine/threonine kinase activity | 78 | 1.32E-10 | 5.52E-08 |
| 10 | GO:0050730 | regulation of peptidyl-tyrosine phosphorylation | 67 | 2.28E-10 | 8.60E-08 |
| 11 | GO:0071902 | positive regulation of protein serine/threonine kinase activity | 57 | 6.73E-10 | 2.30E-07 |
| 12 | GO:0050890 | cognition | 96 | 7.91E-10 | 2.48E-07 |
| 13 | GO:0051897 | positive regulation of phosphatidylinositol 3-kinase/protein kinase B signal transduction | 63 | 1.79E-09 | 5.19E-07 |
| 14 | GO:0042509 | regulation of tyrosine phosphorylation of STAT protein | 27 | 2.64E-09 | 7.09E-07 |
| 15 | GO:0006885 | regulation of pH | 37 | 3.01E-09 | 7.55E-07 |
| 16 | GO:0009581 | detection of external stimulus | 33 | 3.25E-09 | 7.64E-07 |
| 17 | GO:0010638 | positive regulation of organelle organization | 84 | 4.30E-09 | 9.53E-07 |
| 18 | GO:0051452 | intracellular pH reduction | 22 | 5.23E-09 | 1.07E-06 |
| 19 | GO:0007611 | learning or memory | 87 | 5.48E-09 | 1.07E-06 |
| 20 | GO:0044089 | positive regulation of cellular component biogenesis | 82 | 5.69E-09 | 1.07E-06 |
| 21 | GO:0097401 | synaptic vesicle lumen acidification | 14 | 6.89E-09 | 1.23E-06 |
| 22 | GO:0042531 | positive regulation of tyrosine phosphorylation of STAT protein | 26 | 8.40E-09 | 1.44E-06 |
| 23 | GO:0009582 | detection of abiotic stimulus | 34 | 9.27E-09 | 1.49E-06 |
| 24 | GO:0030641 | regulation of cellular pH | 32 | 9.65E-09 | 1.49E-06 |
| 25 | GO:0009612 | response to mechanical stimulus | 78 | 9.89E-09 | 1.49E-06 |

**eTable 4. Gene enrichment results. Gene ontology biological processes categories implicated in bipolar disorder treatment-based scores using over-representation analysis (false discovery rate (FDR) corrected p < 0.05).**

| | GOcategory | Description | NumGenes | Pval | Pval_corr |
|---|---|---|---|---|---|
| 1 | GO:0060078 | regulation of postsynaptic membrane potential | 72 | 0 | 0 |
| 2 | GO:0098664 | G protein-coupled serotonin receptor signaling pathway | 21 | 0 | 0 |
| 3 | GO:0007187 | G protein-coupled receptor signaling pathway, coupled to cyclic nucleotide second messenger | 45 | 0 | 0 |
| 4 | GO:0007188 | adenylate cyclase-modulating G protein-coupled receptor signaling pathway | 92 | 0 | 5.00E-15 |
| 5 | GO:0007193 | adenylate cyclase-inhibiting G protein-coupled receptor signaling pathway | 43 | 0 | 2.26E-13 |
| 6 | GO:0071880 | adenylate cyclase-activating adrenergic receptor signaling pathway | 12 | 1.00E-15 | 3.17E-13 |
| 7 | GO:0071875 | adrenergic receptor signaling pathway | 13 | 7.00E-15 | 3.37E-12 |
| 8 | GO:0051932 | synaptic transmission, GABAergic | 11 | 1.00E-14 | 4.68E-12 |
| 9 | GO:0007214 | gamma-aminobutyric acid signaling pathway | 18 | 1.80E-13 | 7.21E-11 |
| 10 | GO:0001508 | action potential | 45 | 2.67E-13 | 9.64E-11 |
| 11 | GO:0086010 | membrane depolarization during action potential | 26 | 3.04E-13 | 9.96E-11 |
| 12 | GO:0051899 | membrane depolarization | 42 | 8.83E-13 | 2.66E-10 |
| 13 | GO:0007210 | serotonin receptor signaling pathway | 9 | 3.90E-12 | 1.08E-09 |
| 14 | GO:0095500 | acetylcholine receptor signaling pathway | 12 | 1.21E-11 | 3.12E-09 |
| 15 | GO:0098661 | inorganic anion transmembrane transport | 37 | 1.56E-11 | 3.51E-09 |
| 16 | GO:1902476 | chloride transmembrane transport | 37 | 1.56E-11 | 3.51E-09 |
| 17 | GO:0006821 | chloride transport | 38 | 2.57E-11 | 5.44E-09 |
| 18 | GO:0007197 | adenylate cyclase-inhibiting G protein-coupled acetylcholine receptor signaling pathway | 8 | 7.46E-11 | 1.42E-08 |
| 19 | GO:0007213 | G protein-coupled acetylcholine receptor signaling pathway | 8 | 7.46E-11 | 1.42E-08 |
| 20 | GO:0098656 | monoatomic anion transmembrane transport | 41 | 1.03E-10 | 1.86E-08 |
| 21 | GO:0007200 | phospholipase C-activating G protein-coupled receptor signaling pathway | 36 | 1.36E-10 | 2.34E-08 |
| 22 | GO:0098926 | postsynaptic signal transduction | 14 | 1.67E-10 | 2.74E-08 |
| 23 | GO:0006820 | monoatomic anion transport | 43 | 2.42E-10 | 3.79E-08 |
| 24 | GO:0015698 | inorganic anion transport | 44 | 3.62E-10 | 5.44E-08 |
| 25 | GO:0019228 | neuronal action potential | 23 | 4.22E-10 | 6.09E-08 |

**eTable 5. Gene enrichment results. Gene ontology biological processes categories implicated in bipolar disorder GWAS-based scores using over-representation analysis (false discovery rate (FDR) corrected p < 0.05).**

| | GOcategory | Description | NumGenes | Pval | Pval_corr |
|---|---|---|---|---|---|
| 1 | GO:0070588 | calcium ion transmembrane transport | 90 | 1.28E-11 | 4.83E-08 |
| 2 | GO:0032409 | regulation of transporter activity | 85 | 2.09E-10 | 3.94E-07 |
| 3 | GO:0022898 | regulation of transmembrane transporter activity | 78 | 4.14E-10 | 4.72E-07 |
| 4 | GO:0043123 | positive regulation of canonical NF-kappaB signal transduction | 46 | 5.01E-10 | 4.72E-07 |
| 5 | GO:0097553 | calcium ion transmembrane import into cytosol | 48 | 1.34E-09 | 1.01E-06 |
| 6 | GO:0045862 | positive regulation of proteolysis | 74 | 3.78E-09 | 2.37E-06 |
| 7 | GO:0009314 | response to radiation | 95 | 6.39E-09 | 3.07E-06 |
| 8 | GO:0032412 | regulation of monoatomic ion transmembrane transporter activity | 73 | 6.52E-09 | 3.07E-06 |
| 9 | GO:0050806 | positive regulation of synaptic transmission | 51 | 1.24E-08 | 5.18E-06 |
| 10 | GO:0003012 | muscle system process | 96 | 1.44E-08 | 5.27E-06 |
| 11 | GO:0009416 | response to light stimulus | 77 | 1.54E-08 | 5.27E-06 |
| 12 | GO:0051924 | regulation of calcium ion transport | 98 | 1.94E-08 | 6.07E-06 |
| 13 | GO:0010952 | positive regulation of peptidase activity | 40 | 2.47E-08 | 7.16E-06 |
| 14 | GO:0043122 | regulation of canonical NF-kappaB signal transduction | 55 | 4.39E-08 | 1.18E-05 |
| 15 | GO:0051966 | regulation of synaptic transmission, glutamatergic | 38 | 8.86E-08 | 2.21E-05 |
| 16 | GO:0051668 | localization within membrane | 68 | 9.38E-08 | 2.21E-05 |
| 17 | GO:0051092 | positive regulation of NF-kappaB transcription factor activity | 41 | 1.09E-07 | 2.42E-05 |
| 18 | GO:0048167 | regulation of synaptic plasticity | 56 | 1.32E-07 | 2.77E-05 |
| 19 | GO:0009612 | response to mechanical stimulus | 78 | 1.47E-07 | 2.91E-05 |
| 20 | GO:2001056 | positive regulation of cysteine-type endopeptidase activity | 30 | 1.55E-07 | 2.92E-05 |
| 21 | GO:0010950 | positive regulation of endopeptidase activity | 37 | 1.67E-07 | 2.99E-05 |
| 22 | GO:0002831 | regulation of response to biotic stimulus | 98 | 2.12E-07 | 3.62E-05 |
| 23 | GO:0097190 | apoptotic signaling pathway | 77 | 2.40E-07 | 3.94E-05 |
| 24 | GO:1903169 | regulation of calcium ion transmembrane transport | 68 | 3.85E-07 | 6.04E-05 |
| 25 | GO:0035249 | synaptic transmission, glutamatergic | 20 | 4.48E-07 | 6.75E-05 |

**eTable 6. Gene enrichment results. Gene ontology biological processes categories implicated in major depression treatment-based scores using over-representation analysis (false discovery rate (FDR) corrected $p < 0.05$).**

| | GOcategory | Description | NumGenes | Pval | Pval_corr |
|---|---|---|---|---|---|
| 1 | GO:0098664 | G protein-coupled serotonin receptor signaling pathway | 21 | 0 | 0 |
| 2 | GO:0007187 | G protein-coupled receptor signaling pathway, coupled to cyclic nucleotide second messenger | 45 | 0 | 0 |
| 3 | GO:0060078 | regulation of postsynaptic membrane potential | 72 | 0 | 0 |
| 4 | GO:0071880 | adenylate cyclase-activating adrenergic receptor signaling pathway | 12 | 2.00E-15 | 1.73E-12 |
| 5 | GO:0007188 | adenylate cyclase-modulating G protein-coupled receptor signaling pathway | 92 | 1.20E-14 | 9.01E-12 |
| 6 | GO:0071875 | adrenergic receptor signaling pathway | 13 | 2.40E-14 | 1.42E-11 |
| 7 | GO:0051932 | synaptic transmission, GABAergic | 11 | 3.40E-14 | 1.74E-11 |
| 8 | GO:0007193 | adenylate cyclase-inhibiting G protein-coupled receptor signaling pathway | 43 | 4.70E-14 | 2.10E-11 |
| 9 | GO:0001508 | action potential | 45 | 1.39E-13 | 5.56E-11 |
| 10 | GO:0007214 | gamma-aminobutyric acid signaling pathway | 18 | 7.12E-13 | 2.57E-10 |
| 11 | GO:0086010 | membrane depolarization during action potential | 26 | 1.46E-12 | 4.79E-10 |
| 12 | GO:0007210 | serotonin receptor signaling pathway | 9 | 1.01E-11 | 3.05E-09 |
| 13 | GO:0019228 | neuronal action potential | 23 | 7.27E-11 | 1.90E-08 |
| 14 | GO:0098661 | inorganic anion transmembrane transport | 37 | 7.91E-11 | 1.90E-08 |
| 15 | GO:1902476 | chloride transmembrane transport | 37 | 7.91E-11 | 1.90E-08 |
| 16 | GO:0006821 | chloride transport | 38 | 1.30E-10 | 2.92E-08 |
| 17 | GO:0007197 | adenylate cyclase-inhibiting G protein-coupled acetylcholine receptor signaling pathway | 8 | 1.74E-10 | 3.49E-08 |
| 18 | GO:0007213 | G protein-coupled acetylcholine receptor signaling pathway | 8 | 1.74E-10 | 3.49E-08 |
| 19 | GO:0098656 | monoatomic anion transmembrane transport | 41 | 5.12E-10 | 9.72E-08 |
| 20 | GO:0007200 | phospholipase C-activating G protein-coupled receptor signaling pathway | 36 | 6.17E-10 | 1.11E-07 |
| 21 | GO:0006820 | monoatomic anion transport | 43 | 1.18E-09 | 2.03E-07 |
| 22 | GO:0034776 | response to histamine | 9 | 1.49E-09 | 2.44E-07 |
| 23 | GO:0015698 | inorganic anion transport | 44 | 1.76E-09 | 2.65E-07 |
| 24 | GO:0030534 | adult behavior | 44 | 1.76E-09 | 2.65E-07 |
| 25 | GO:0095500 | acetylcholine receptor signaling pathway | 12 | 1.90E-09 | 2.74E-07 |

**eTable 7. Gene enrichment results. Gene ontology biological processes categories implicated in major depression GWAS-based scores using over-representation analysis (false discovery rate (FDR) corrected p < 0.05).**

| | GOcategory | Description | NumGenes | Pval | Pval_corr |
|---|---|---|---|---|---|
| 1 | GO:0022904 | respiratory electron transport chain | 53 | 4.79E-12 | 1.80E-08 |
| 2 | GO:0019646 | aerobic electron transport chain | 47 | 1.30E-10 | 2.45E-07 |
| 3 | GO:0009060 | aerobic respiration | 54 | 7.89E-10 | 8.33E-07 |
| 4 | GO:0006974 | DNA damage response | 83 | 8.85E-10 | 8.33E-07 |
| 5 | GO:0006120 | mitochondrial electron transport, NADH to ubiquinone | 34 | 5.59E-09 | 2.92E-06 |
| 6 | GO:0033108 | mitochondrial respiratory chain complex assembly | 34 | 5.59E-09 | 2.92E-06 |
| 7 | GO:0032774 | RNA biosynthetic process | 50 | 6.11E-09 | 2.92E-06 |
| 8 | GO:1903047 | mitotic cell cycle process | 63 | 6.20E-09 | 2.92E-06 |
| 9 | GO:0044089 | positive regulation of cellular component biogenesis | 82 | 7.21E-09 | 3.01E-06 |
| 10 | GO:0006259 | DNA metabolic process | 74 | 9.05E-09 | 3.41E-06 |
| 11 | GO:2001233 | regulation of apoptotic signaling pathway | 98 | 1.50E-08 | 5.13E-06 |
| 12 | GO:0051966 | regulation of synaptic transmission, glutamatergic | 38 | 1.74E-08 | 5.38E-06 |
| 13 | GO:0000122 | negative regulation of transcription by RNA polymerase II | 87 | 1.86E-08 | 5.38E-06 |
| 14 | GO:0022402 | cell cycle process | 93 | 2.80E-08 | 7.14E-06 |
| 15 | GO:0010257 | NADH dehydrogenase complex assembly | 31 | 3.04E-08 | 7.14E-06 |
| 16 | GO:0032981 | mitochondrial respiratory chain complex I assembly | 31 | 3.04E-08 | 7.14E-06 |
| 17 | GO:0006351 | DNA-templated transcription | 42 | 3.22E-08 | 7.14E-06 |
| 18 | GO:0042113 | B cell activation | 46 | 4.61E-08 | 9.65E-06 |
| 19 | GO:0034330 | cell junction organization | 77 | 6.12E-08 | 1.21E-05 |
| 20 | GO:0045333 | cellular respiration | 57 | 1.03E-07 | 1.93E-05 |
| 21 | GO:0006366 | transcription by RNA polymerase II | 34 | 1.50E-07 | 2.69E-05 |
| 22 | GO:0051098 | regulation of binding | 56 | 1.62E-07 | 2.78E-05 |
| 23 | GO:0009887 | animal organ morphogenesis | 74 | 2.15E-07 | 3.52E-05 |
| 24 | GO:0050806 | positive regulation of synaptic transmission | 51 | 2.58E-07 | 4.05E-05 |
| 25 | GO:0010564 | regulation of cell cycle process | 97 | 3.00E-07 | 4.36E-05 |

**eTable 8. Gene enrichment results. Gene ontology biological processes categories implicated in schizophrenia treatment-based scores using over-representation analysis (false discovery rate (FDR) corrected p < 0.05).**

| | GOcategory | Description | NumGenes | Pval | Pval_corr |
|---|---|---|---|---|---|
| 1 | GO:0098664 | G protein-coupled serotonin receptor signaling pathway | 21 | 0 | 0 |
| 2 | GO:0007187 | G protein-coupled receptor signaling pathway, coupled to cyclic nucleotide second messenger | 45 | 0 | 0 |
| 3 | GO:0007188 | adenylate cyclase-modulating G protein-coupled receptor signaling pathway | 92 | 0 | 0 |
| 4 | GO:0007193 | adenylate cyclase-inhibiting G protein-coupled receptor signaling pathway | 43 | 0 | 0 |
| 5 | GO:0071880 | adenylate cyclase-activating adrenergic receptor signaling pathway | 12 | 0 | 2.00E-15 |
| 6 | GO:0071875 | adrenergic receptor signaling pathway | 13 | 0 | 1.90E-14 |
| 7 | GO:0060078 | regulation of postsynaptic membrane potential | 72 | 9.00E-15 | 4.41E-12 |
| 8 | GO:0007210 | serotonin receptor signaling pathway | 9 | 7.70E-14 | 3.46E-11 |
| 9 | GO:0007200 | phospholipase C-activating G protein-coupled receptor signaling pathway | 36 | 2.30E-13 | 9.22E-11 |
| 10 | GO:0007197 | adenylate cyclase-inhibiting G protein-coupled acetylcholine receptor signaling pathway | 8 | 2.32E-12 | 6.75E-10 |
| 11 | GO:0007213 | G protein-coupled acetylcholine receptor signaling pathway | 8 | 2.32E-12 | 6.75E-10 |
| 12 | GO:0035150 | regulation of tube size | 65 | 2.62E-12 | 6.75E-10 |
| 13 | GO:0035296 | regulation of tube diameter | 65 | 2.62E-12 | 6.75E-10 |
| 14 | GO:0097746 | blood vessel diameter maintenance | 65 | 2.62E-12 | 6.75E-10 |
| 15 | GO:0007214 | gamma-aminobutyric acid signaling pathway | 18 | 8.64E-11 | 2.08E-08 |
| 16 | GO:0003018 | vascular process in circulatory system | 82 | 1.91E-10 | 4.31E-08 |
| 17 | GO:0003013 | circulatory system process | 94 | 2.54E-10 | 5.38E-08 |
| 18 | GO:0042310 | vasoconstriction | 16 | 7.12E-10 | 1.43E-07 |
| 19 | GO:0095500 | acetylcholine receptor signaling pathway | 12 | 1.02E-09 | 1.93E-07 |
| 20 | GO:0090066 | regulation of anatomical structure size | 96 | 3.01E-09 | 5.43E-07 |
| 21 | GO:0098926 | postsynaptic signal transduction | 14 | 5.82E-09 | 9.99E-07 |
| 22 | GO:0007189 | adenylate cyclase-activating G protein-coupled receptor signaling pathway | 51 | 1.26E-08 | 2.06E-06 |
| 23 | GO:0051932 | synaptic transmission, GABAergic | 11 | 2.02E-08 | 3.17E-06 |
| 24 | GO:0007198 | adenylate cyclase-inhibiting serotonin receptor signaling pathway | 5 | 5.89E-08 | 8.85E-06 |
| 25 | GO:0007212 | dopamine receptor signaling pathway | 13 | 9.91E-08 | 1.43E-05 |

**eTable 9. Gene enrichment results. Gene ontology biological processes categories implicated in schizophrenia GWAS-based scores using over-representation analysis (false discovery rate (FDR) corrected p < 0.05).**

| | GOcategory | Description | NumGenes | Pval | Pval_corr |
|---|---|---|---|---|---|
| 1 | GO:2001233 | regulation of apoptotic signaling pathway | 98 | 1.72E-09 | 6.47E-06 |
| 2 | GO:2001234 | negative regulation of apoptotic signaling pathway | 67 | 3.65E-07 | 6.87E-04 |
| 3 | GO:0009896 | positive regulation of catabolic process | 98 | 8.71E-07 | 8.51E-04 |
| 4 | GO:0070588 | calcium ion transmembrane transport | 90 | 9.05E-07 | 8.51E-04 |
| 5 | GO:0010506 | regulation of autophagy | 88 | 1.43E-06 | 1.04E-03 |
| 6 | GO:0016241 | regulation of macroautophagy | 50 | 1.67E-06 | 1.04E-03 |
| 7 | GO:0032409 | regulation of transporter activity | 85 | 2.83E-06 | 1.34E-03 |
| 8 | GO:0097553 | calcium ion transmembrane import into cytosol | 48 | 2.85E-06 | 1.34E-03 |
| 9 | GO:0019646 | aerobic electron transport chain | 47 | 3.74E-06 | 1.56E-03 |
| 10 | GO:0051098 | regulation of binding | 56 | 6.06E-06 | 2.28E-03 |
| 11 | GO:0007005 | mitochondrion organization | 43 | 1.10E-05 | 3.53E-03 |
| 12 | GO:0031331 | positive regulation of cellular catabolic process | 71 | 1.13E-05 | 3.53E-03 |
| 13 | GO:0022898 | regulation of transmembrane transporter activity | 78 | 1.36E-05 | 3.94E-03 |
| 14 | GO:0034330 | cell junction organization | 77 | 1.70E-05 | 3.99E-03 |
| 15 | GO:0070663 | regulation of leukocyte proliferation | 77 | 1.70E-05 | 3.99E-03 |
| 16 | GO:1904951 | positive regulation of establishment of protein localization | 77 | 1.70E-05 | 3.99E-03 |
| 17 | GO:0000302 | response to reactive oxygen species | 76 | 2.12E-05 | 4.41E-03 |
| 18 | GO:0032944 | regulation of mononuclear cell proliferation | 68 | 2.24E-05 | 4.41E-03 |
| 19 | GO:0050670 | regulation of lymphocyte proliferation | 68 | 2.24E-05 | 4.41E-03 |
| 20 | GO:0019221 | cytokine-mediated signaling pathway | 97 | 2.34E-05 | 4.41E-03 |
| 21 | GO:0032774 | RNA biosynthetic process | 50 | 2.74E-05 | 4.88E-03 |
| 22 | GO:0080135 | regulation of cellular response to stress | 82 | 2.85E-05 | 4.88E-03 |
| 23 | GO:0032412 | regulation of monoatomic ion transmembrane transporter activity | 73 | 4.08E-05 | 6.68E-03 |
| 24 | GO:0031400 | negative regulation of protein modification process | 86 | 5.39E-05 | 7.94E-03 |
| 25 | GO:0032386 | regulation of intracellular transport | 37 | 5.48E-05 | 7.94E-03 |
